# Remote working in mental health services: a rapid umbrella review of pre-COVID-19 literature

**DOI:** 10.1101/2020.11.30.20240721

**Authors:** Phoebe Barnett, Lucy Goulding, Cecilia Casetta, Harriet Jordan, Luke Sheridan-Rains, Thomas Steare, Julie Williams, Lisa Wood, Fiona Gaughran, Sonia Johnson

## Abstract

**Background:** Tele-mental health care has been rapidly adopted to maintain services during the pandemic, and there is now substantial interest in its future role. Service planning and policy making for recovery from the pandemic and beyond should draw not only on COVID-19 experiences, but also on the substantial research evidence accumulated prior to this.

**Aims:** to conduct an umbrella review of systematic reviews of research literature and evidence-based guidance on remote working in mental health, including both qualitative and quantitative literature.

**Method:** Three databases were searched between January 2010 and August 2020 for systematic reviews meeting pre-defined criteria. Reviews retrieved were independently screened and those meeting inclusion criteria were synthesised and assessed for risk of bias. Narrative synthesis was used to report findings

**Results:** Nineteen systematic reviews met inclusion criteria. Fifteen examined clinical effectiveness, eight reported on aspects of tele-mental health implementation, ten reported on acceptability to service users and clinicians, two on cost-effectiveness and one on guidance. Most reviews were assessed as low quality. Findings suggested that video-based communication could be as effective and acceptable as face-face formats, at least in the short-term. Evidence was lacking on extent of digital exclusion and how it can be overcome, or on significant context such as children and young people and inpatient settings.

**Conclusions:** This umbrella review suggests that tele-mental health has potential to be an effective and acceptable form of service delivery. However, we found limited evidence on impacts of large-scale implementation across catchment areas. Combining previous evidence and COVID-19 experiences may allow realistic planning for future tele-mental health implementation.

## Introduction

Mental health care and treatment utilising remote technologies such as video or phone (tele-mental health) has become an important tool in recent months, taking a central role internationally in maintaining mental health services during the COVID-19 pandemic (1). Policy makers and mental health professionals, along with mental health service users now express interest in continuing some use of these technologies long-term, even in the absence of pandemic-related social distancing requirements (1-3). Potential benefits of remote technologies extend beyond adaptation to government social distancing guidelines, allowing the efficiency and flexibility of mental health services to be maximised. The mobilisation of tele-mental health during the pandemic has happened largely ad-hoc, achieving remarkably rapid but highly variable implementation. This emergency response has largely occurred without systematic reference to previous literature. In order to plan effective and acceptable deployment of tele-mental health beyond the pandemic, it is crucial that we now take stock of all relevant evidence regarding potential impacts, challenges and outcomes of widespread remote technology utilisation and identify key mechanisms for its acceptable integration into routine care, (4).

Tele-mental health has a number of potential benefits that make it of significant interest to service providers not only during the pandemic, but also longer-term: For service users across a range of populations, settings and conditions (5), potential benefits include convenience and improved accessibility, particularly where issues such as physical mobility difficulties, anxiety, or paranoia impede face-to-face contacts (1). Potential advantages for staff include reduced environmental impact, greater convenience and opportunities for home working and ease of effective communication within and between mental health teams (2). Although some have argued that problems with building of rapport (6), and privacy or safety concerns may hinder implementation of remote care, service users have been found to report such apprehensions less than clinicians (7). Several studies have also suggested that tele-mental health may be more cost effective than face-to-face delivery. (7)

Despite potential benefits and efficiencies, and a substantial body of relevant research, implementation of remote working remained very limited in most countries prior to the pandemic, and substantial implementation barriers have been observed (8), along with potential for inequalities to be exacerbated. Digital exclusion is an important concern regarding service users without the necessary skills, equipment and monetary resources to access online treatment, with this most marked in more marginalised groups such as people from BAME and low-SES backgrounds, and loss of privacy and deterioration in therapeutic relationships are further risks (1, 9-11). Staff participation is also impeded by technological and environmental difficulties, and they express reservations regarding quality of assessments, deterioration of therapeutic relationships, and limitations in the extent to which physical as well as mental health is attended to (8, 10, 11).

Thus, potential benefits and disadvantages of tele-mental health are finely balanced. Risks of longer-term roll-out of remote working without close attention to intended and unintended consequences include digital exclusion of some of those already most disadvantaged and decline in quality of care and potentially of outcomes. One source with potential to inform policy makers and service planners in their future tele-mental health strategies is the substantial body of research studies published before the pandemic. We have therefore aimed to provide a rapid summary of the existing literature on the effectiveness, cost-effectiveness, barriers and facilitators for implementation, acceptability and reach of remote interventions for assessment and treatment of mental health problems. Our objective was to identify, appraise and synthesise systematic reviews of literature and guidance on remote working in mental health, including qualitative and quantitative outcomes using “umbrella review” or “review of reviews” methodology. Umbrella reviews are useful when the evidence base is broad, and are useful in summarising a broad evidence base in order to inform policy (12). It is hoped that the results may help to illuminate the benefits and remaining challenges when implementing telehealth technologies during the remainder of the pandemic and in the perhaps permanently changed reality that follows.

## Method

A rapid umbrella review was conducted, guided by the World Health Organisation (WHO) practical guide for *Rapid Reviews to Strengthen Health Policy and Systems* (13) and adhering to Preferred Reporting Items for Systematic reviews and Meta-Analyses (PRISMA) guidelines (14) and umbrella review guidance (15). In line with agreed rapid review methodology, our aim in this review was to provide a timely but robust answer to the research question, through accelerating some aspects of the systematic review process while maintaining transparency and protocol-driven decision making throughout (13). The protocol was prospectively registered on PROSPERO (CRD42020208085).

### Search strategy and selection criteria

The search strategy implemented a combination of keyword and subject heading searches across PsycINFO (01/01/2010-26/08/2020), PubMed (01/01/2010-26/08/2020) and the Cochrane Database of Systematic Reviews (01/01/2010-26/08/2020). The full search strategy is available in Appendix 1. We included systematic reviews meeting the following criteria:

#### Population

Staff working within the field of mental health, people receiving mental health care or with mental health diagnoses, family members or carers of people receiving mental health care. We included people with dementia, neurodevelopmental disorders and addiction, but excluded people with primary sleep disorders unless combined with another included mental health problem.

#### Interventions

Any form of spoken or written communication carried out between mental health professionals and patients/service users/family members /carers or between mental health professionals using either the internet or the telephone. We excluded reviews of digital interventions where the primary aim of the technology was not to facilitate direct therapeutic contact with a mental health professional: thus, for example we excluded apps and websites delivering assessment or treatment in a digital format.

#### Outcomes

Reviews reporting at least one of: implementation outcomes (outcomes relating to the process of care, adherence to intended models, uptake and coverage and barriers and facilitators to implementation), acceptability outcomes (including staff and service user satisfaction, and experiences of the therapeutic relationship and communication), clinical effectiveness, cost effectiveness, or evidence-based guidance for remote working were included. Qualitative and quantitative data were included.

#### Design

Systematic reviews with or without meta-analyses, realist reviews, and qualitative meta-syntheses were included. We considered reviews to be of sufficient quality for inclusion if they searched at least 3 databases, and in line with recommendations for the conduct of systematic reviews for quantitative data (14), quantitative reviews were also required to include appraisal of the quality of included studies.

Due to the rapid nature of the review, we limited our search to reviews published since January 2010 and those available in English language. This was a pragmatic decision taken since studies published prior to 2010 would still be picked up within systematic reviews.

Three reviewers (PB, LG, CC) double screened 10% of titles and abstracts, with disagreements being discussed until consensus was reached. The remaining titles were then independently screened, with studies not meeting inclusion criteria excluded. Full-text articles were subsequently reviewed by five reviewers (PB, TS, LG, CC, LW). A selection of full-texts were double checked to ensure consistency, and any reviews which did not facilitate a straightforward inclusion or exclusion decision were discussed with the wider review group. The search and screening process is depicted in Figure 1.

**Figure 1:**
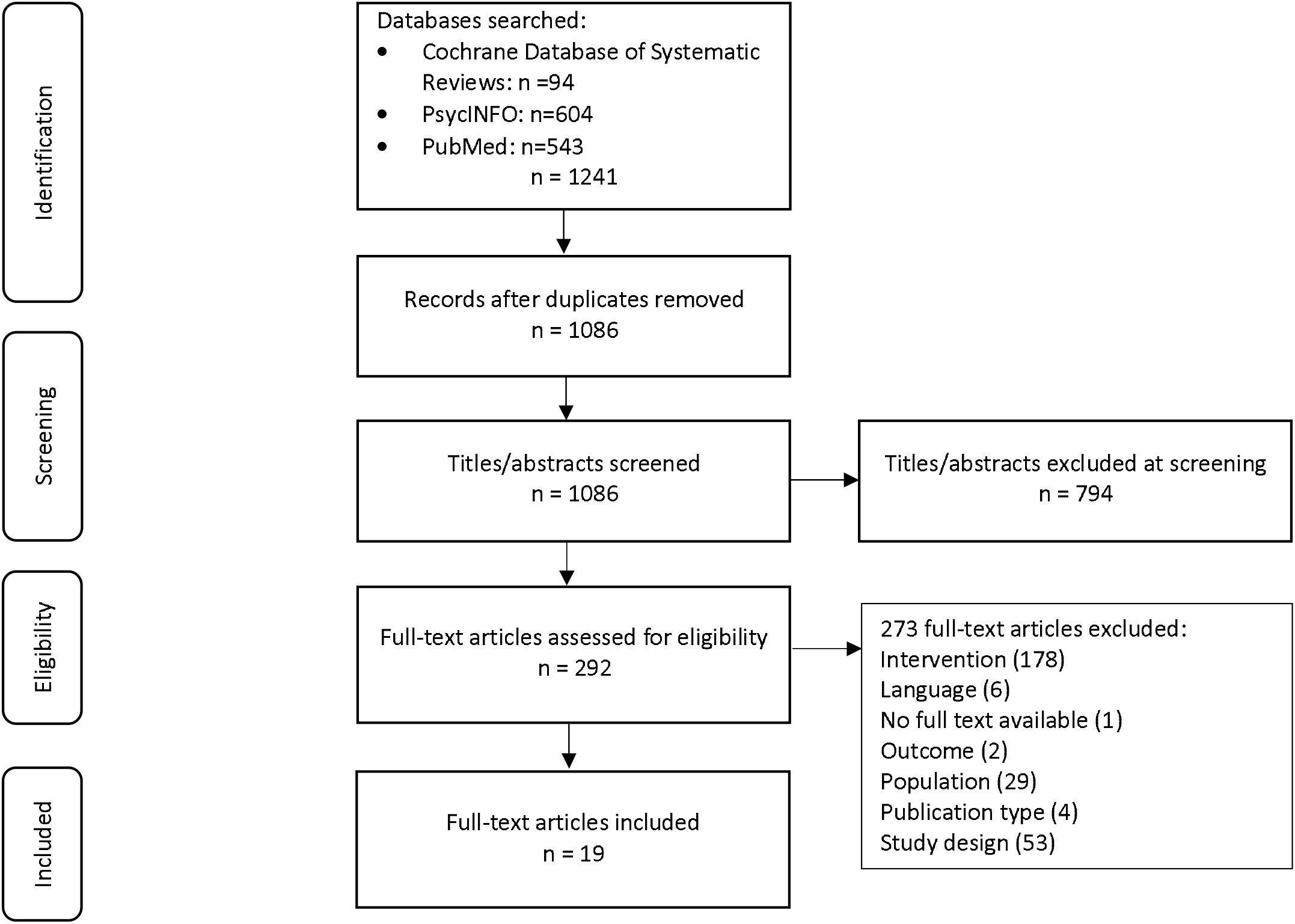
Prisma Diagram.

### Data extraction

Seven reviewers (LG, CC, PB, TS, LSR, JW, HIJ) extracted data from included reviews using an Excel-based form. 10% of extractions were double checked by a second reviewer, and inconsistencies discussed and corrected. Data extracted included: citation details, objectives, type of review, participant details (including gender, ethnicity, age, and mental health diagnosis and staff details where relevant), type(s) of tele-mental health intervention reviewed, setting and context (mental health service, community or inpatient/residential, primary mental health care service), number of databases sourced and searched, date range of database searching, publication date range of studies included in the review informing each outcome of interest, number of included studies, types of studies and country of origin of studies included, instrument used to appraise the primary studies and the rating of their quality, reported clinical, cost-effectiveness and implementation outcomes, method of synthesis/analysis employed to synthesize the evidence, conclusions of the review authors.

### Quality assessment

Quality of each included systematic review was assessed using the Assessment of Multiple Systematic Reviews (AMSTAR2) checklist (16). The checklist was used to give each review an overall rating of quality ranging from high to critically low based on review design weaknesses (16). Study quality was assessed alongside data extraction, and quality ratings are available in Table 1.

**Table 1:**
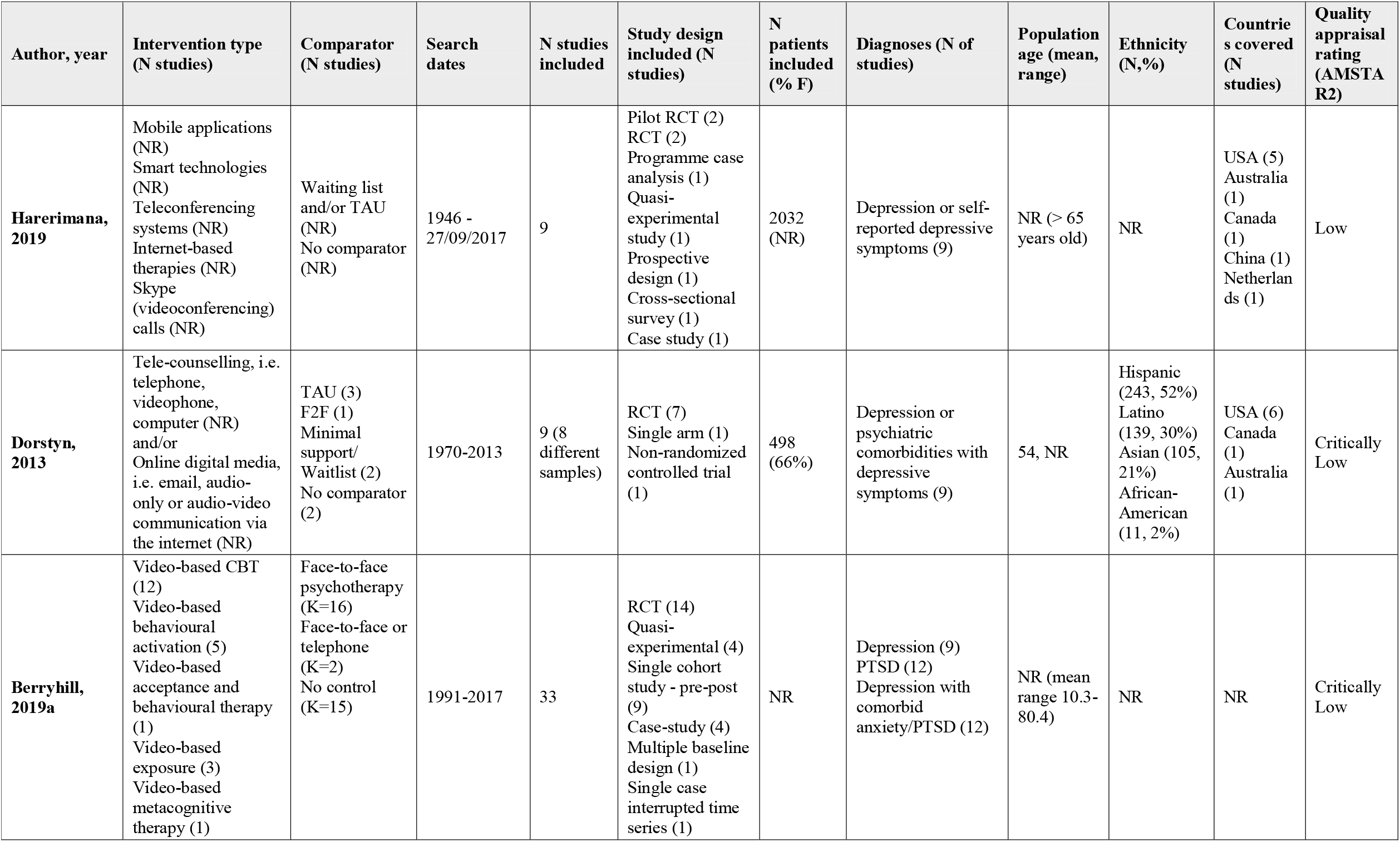
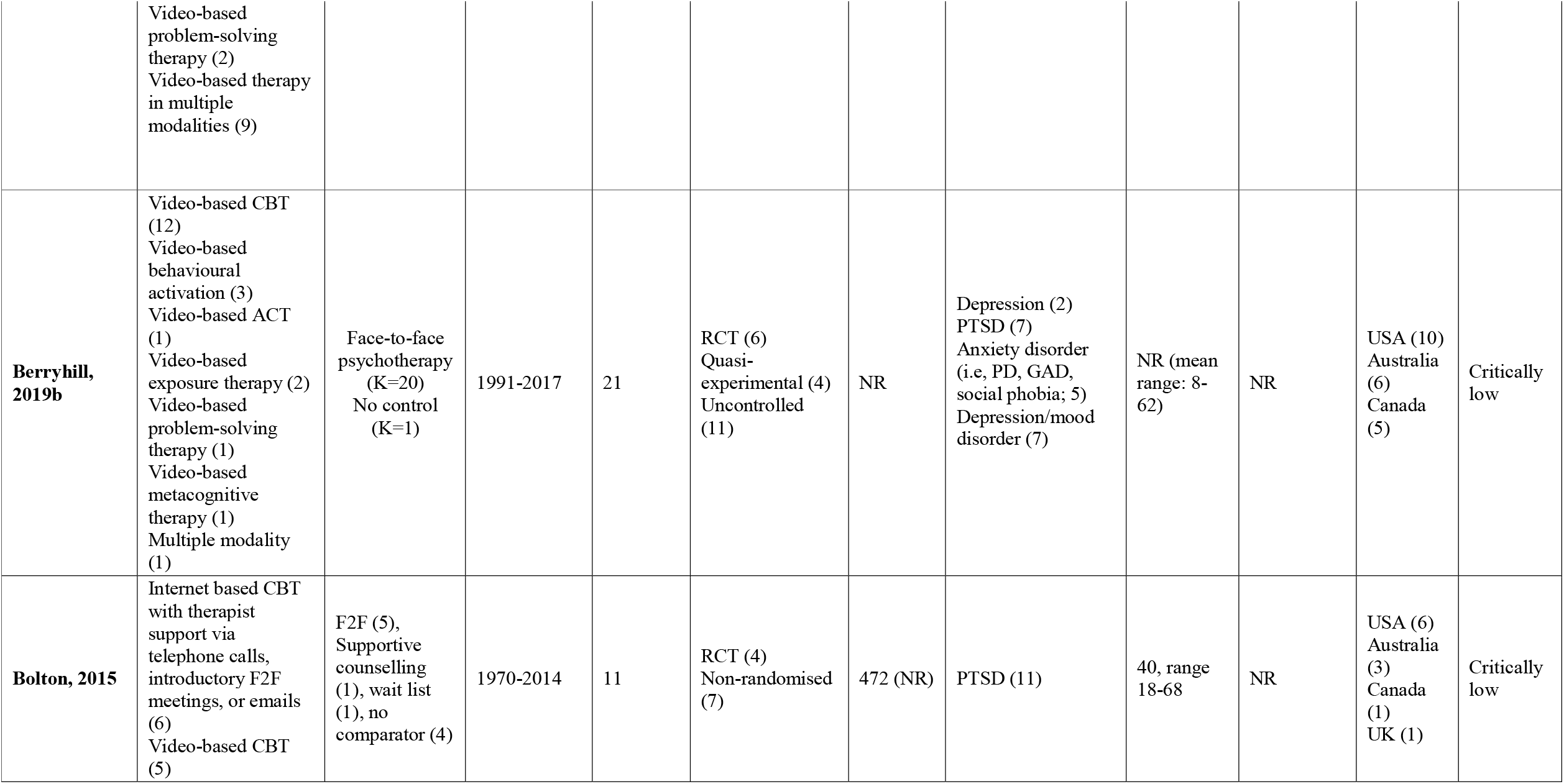
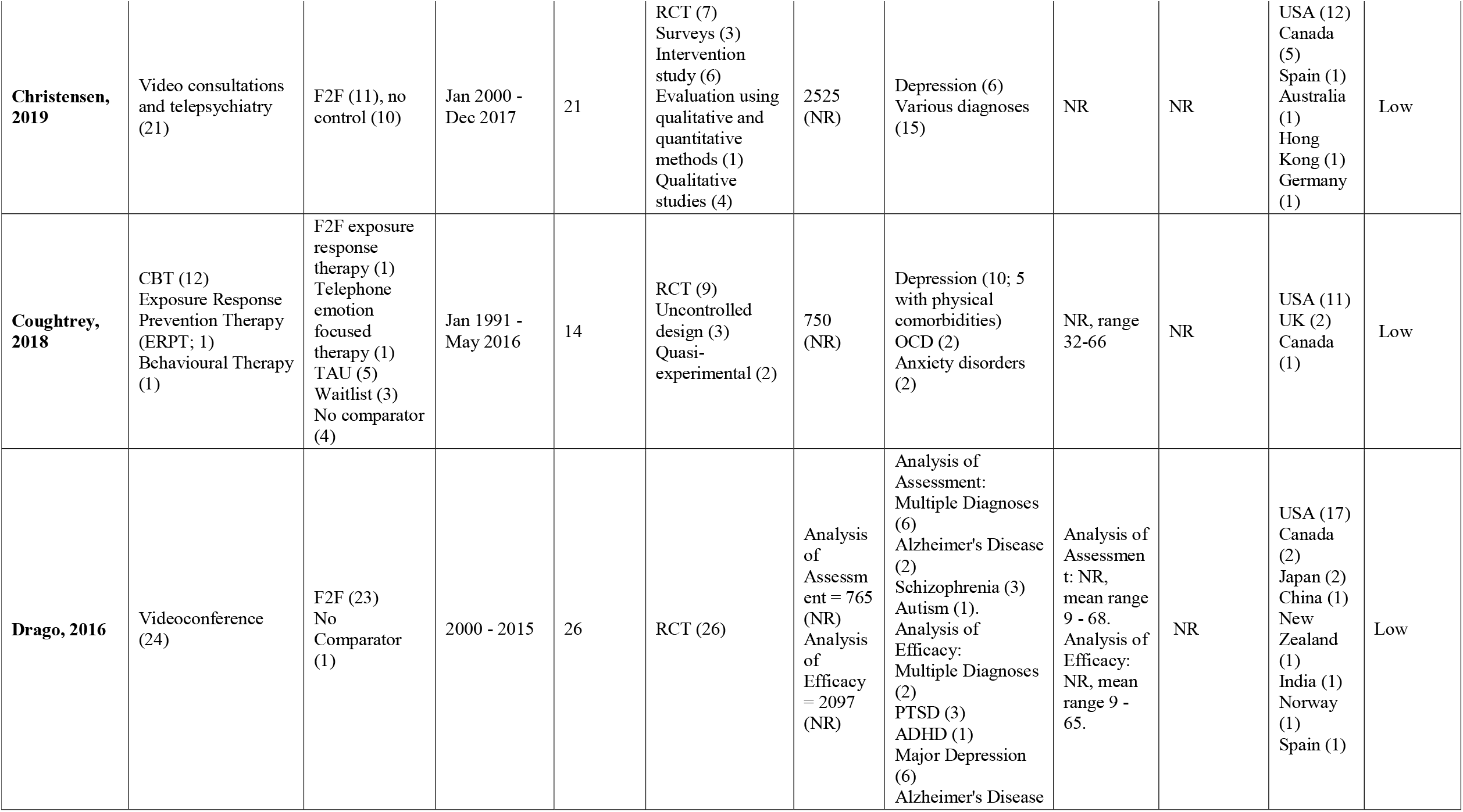
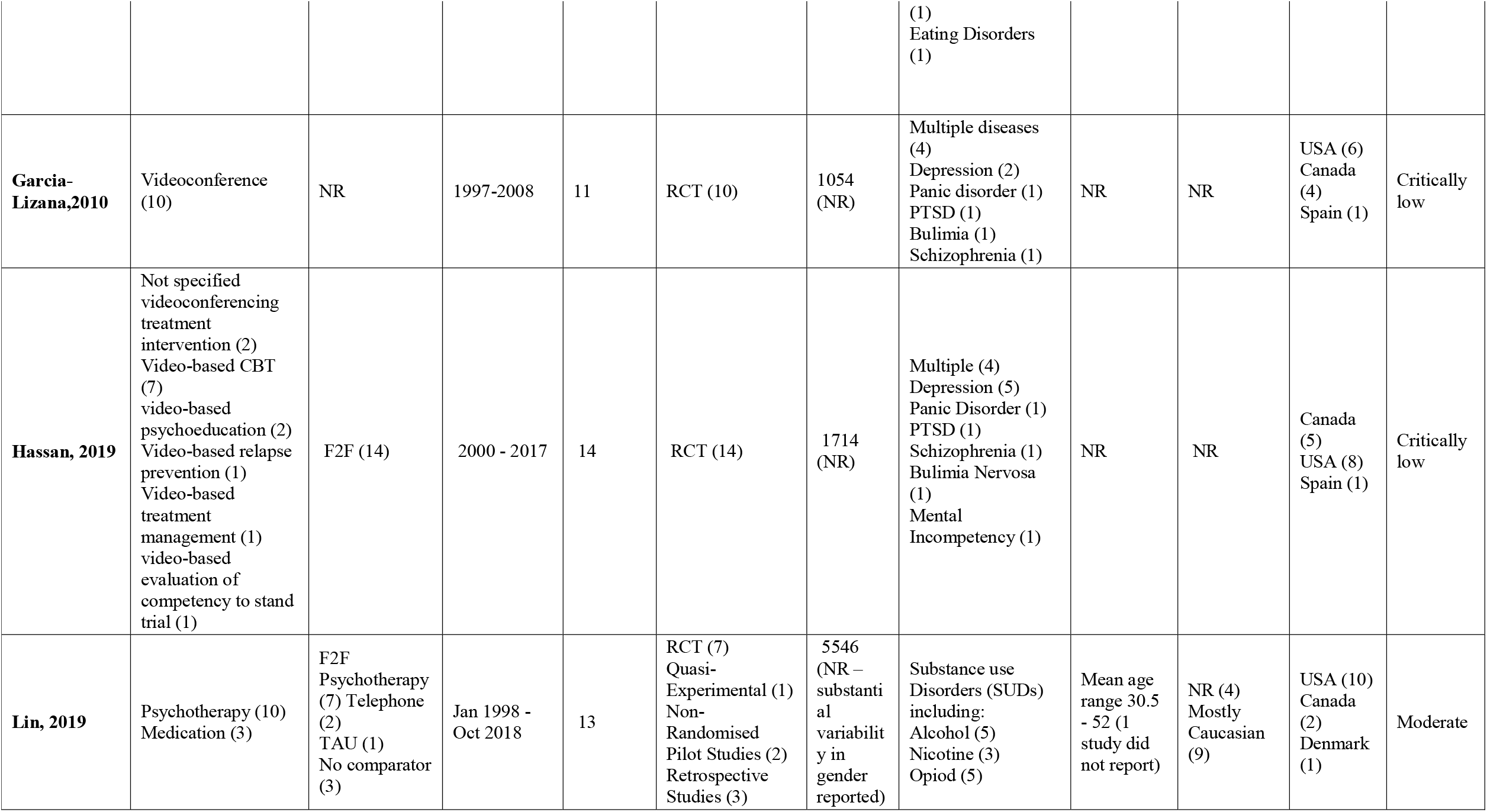
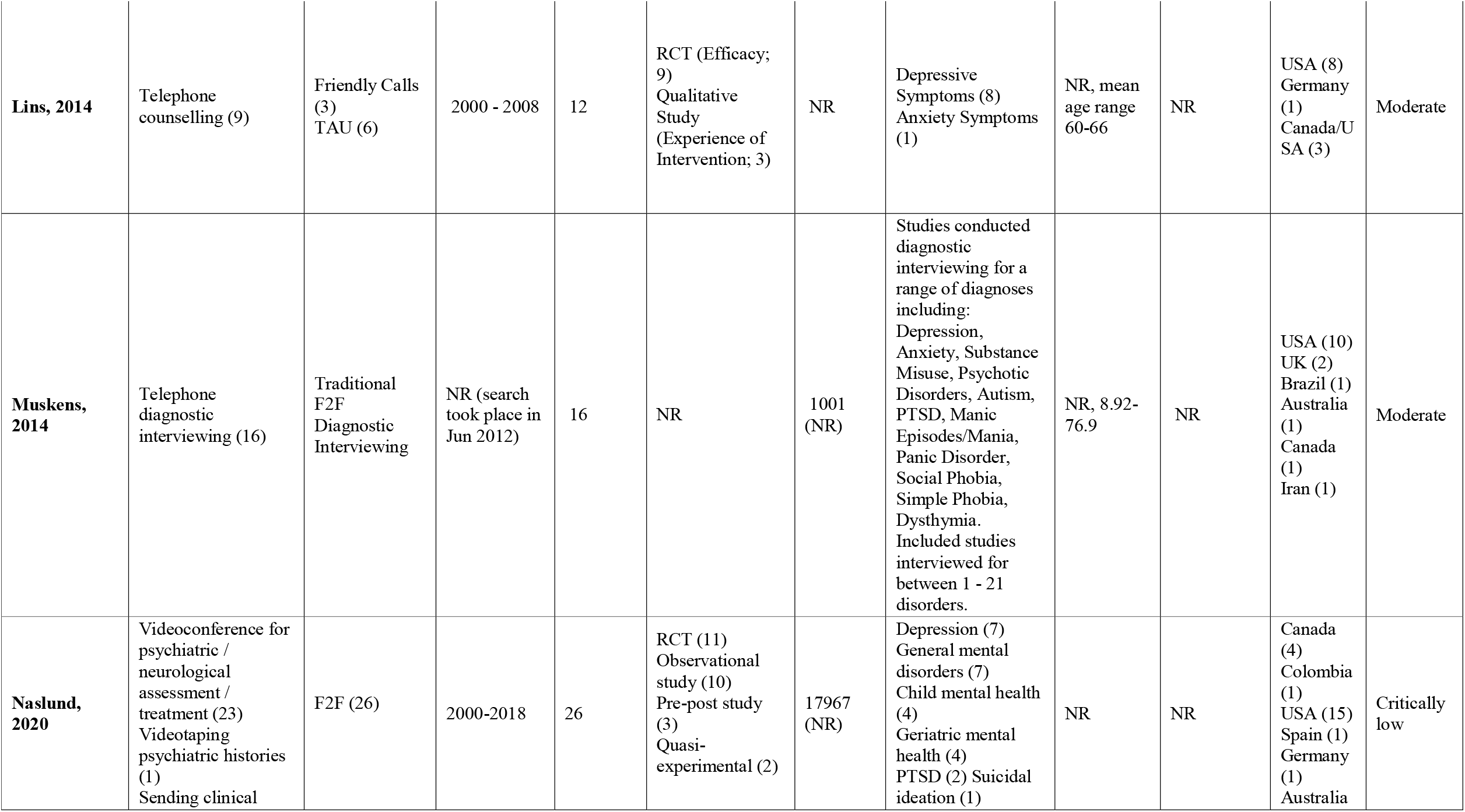
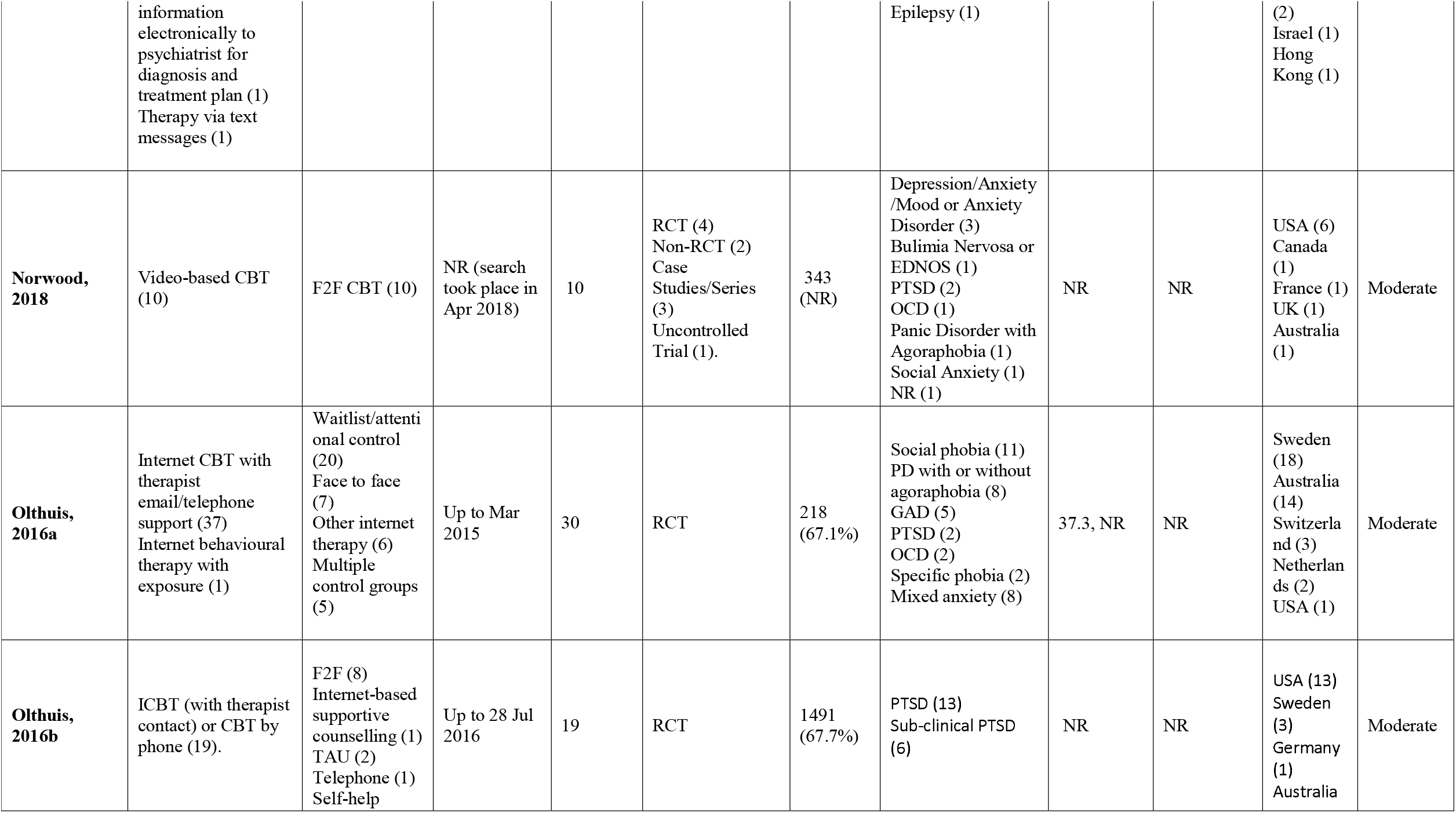
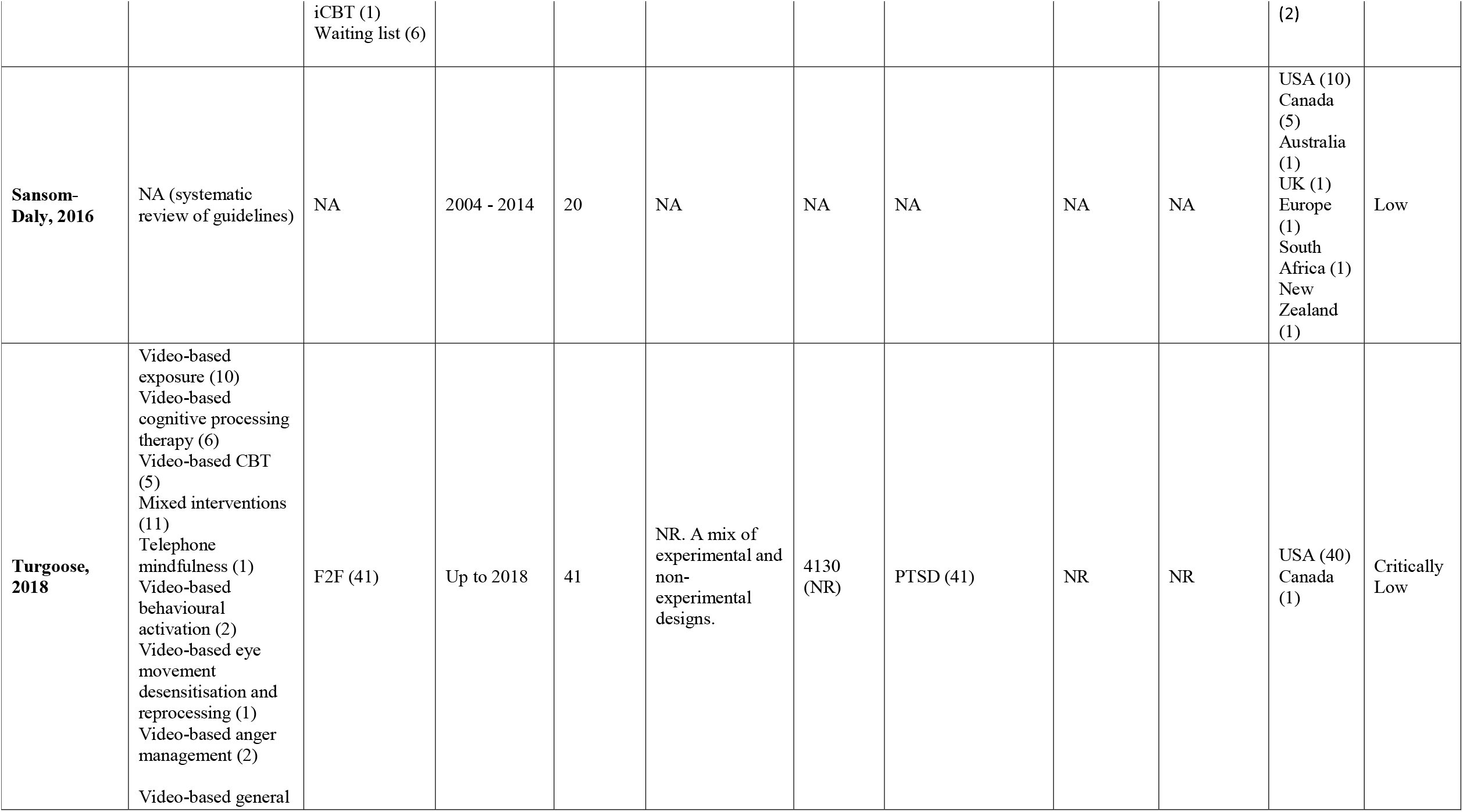
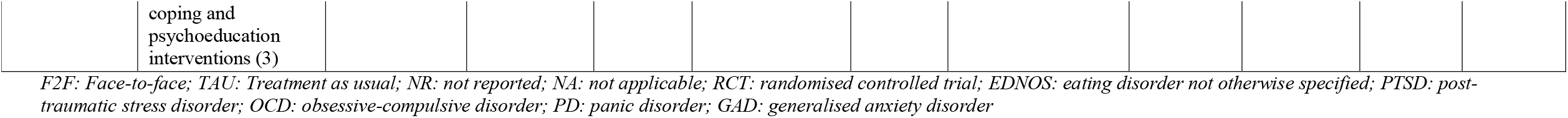
Characteristics of included studies.

### Data synthesis

Heterogeneity in study populations and interventions included in the review, as well as broad inclusion criteria for review study design (e.g. qualitative, quantitative), prevented quantitative pooling of syntheses. As a result, we conducted a narrative synthesis of all interventions and outcomes (17). This allowed a more in-depth consideration of all outcome measures and variations in remote intervention delivery. We grouped reviews by the included population (mental health diagnosis), and further considered the variation in interventions on offer within these subgroups. This was done for each outcome of interest. Most reviews provided a synthesis of multiple intervention types, or failed to adequately differentiate them, making a more thorough comparison across formats impossible.

## Results

The search returned 1,086 reviews, from which 292 potentially relevant full-text articles were identified. Following full text checks, 19 reviews met the inclusion criteria (See Figure 1), reporting on 239 individual studies and 20 guidance documents. Fifteen of the included reviews examined the clinical effectiveness of tele-mental health compared to (a) face-to-face interventions and assessments (K=4), (b)Treatment as usual (K=2) or (c) a variety of comparators including face-to-face, telephone and treatment as usual (K=9). Eight reviews reported on implementation (broadly defined), including process variable, fidelity and uptake of interventions, and ten reviews reported outcomes relating to acceptability, including satisfaction of both service users and clinicians. One review focused specifically on the difference in therapeutic alliance between treatment modalities. Two reviews reported on cost-effectiveness, one on this topic only and the other in combination with clinical effectiveness. One review synthesised international guidance on the conduct of videoconferencing based mental health treatments. Full details of included reviews are available in Table 1. Some primary studies were included in more than one review: 26 studies appeared in two reviews and 27 studies appeared in 3 or more. The remaining 186 studies appeared in only one review. Double-counting of primary studies due to inclusion in multiple reviews contributing to the same outcome was only found for clinical effectiveness outcomes. However, conclusions were similar across reviews, even though no review had all the same studies contributing to each synthesis. Further details of study overlap can be found in Appendix 2.

### Quality of included reviews

Most reviews elicited low confidence on quality appraisal due to multiple study design weaknesses. The most common weaknesses included a lack of explicit statements that a protocol was developed prior to commencement of the review (Explicit statements were reported in two reviews (18, 19)), lack of duplicate study selection (duplicate selection was reported in five reviews (19-23)), no report of excluded studies and reasons for exclusion (exclusions were reported in two reviews, (19, 21)), and no report of sources of funding (sources of funding were reported in three reviews, (21, 24, 25)). Meta-analysis was not performed in the majority of reviews, usually due to heterogeneous data or aims centring around more narrative conclusions such as satisfaction (K=12), but in those that included meta-analysis (19, 21, 25-28), all except two (21, 27) assessed publication bias. The potential impact of risk of bias was only assessed in two reviews performing meta-analysis (21, 27), but all reviews performing meta-analysis used appropriate statistical methods for combining results. The reviews eliciting higher confidence (moderate) were the two Cochrane reviews (21, 28). Quality ratings of reviews are available in Table 1, and full details of quality assessments are available in Appendix 3

### Clinical outcomes

Clinical outcomes were reported in 15 reviews (18-21, 24-34). Across all patient populations, including patients with anxiety (K=3), PTSD (K=2), depression (K=4) (including in ethnic minorities (K=1)(31) and older adults (K=1)(18)), substance use disorders (K=1) and multiple disorders (K=4), videoconferencing interventions were reported to result in significant reductions in symptom severity, with outcomes comparable to face-to-face controls where these were included. Telephone based interventions tended to report similar significant reductions in symptom severity. However, the review of telephone interventions with older adults with depression (18) reported more mixed findings: reductions were reported in emergency room and hospital visits in one study, and in depression in another, but a third study suggested that telephone interventions did not add to benefit from a web-only intervention. n. Follow-up treatment gains were less widely reported and conclusions were mixed across reviews. While maintenance of improvements was found at follow up assessments in two reviews regarding video-based tele-therapy (27, 34) and another regarding telephone-based therapy (24), two other reviews reported that videoconference interventions may show less longevity in maintenance of effects than face-to-face interventions (26, 31). A final review of mixed modality remote interventions suggested that while inferior to face to face formats at shorter term follow up, remote interventions may be more beneficial than face-to-face at longer follow-ups (36 months) (18). Further details on clinical outcomes are available in Table 2.

**Table 2:**
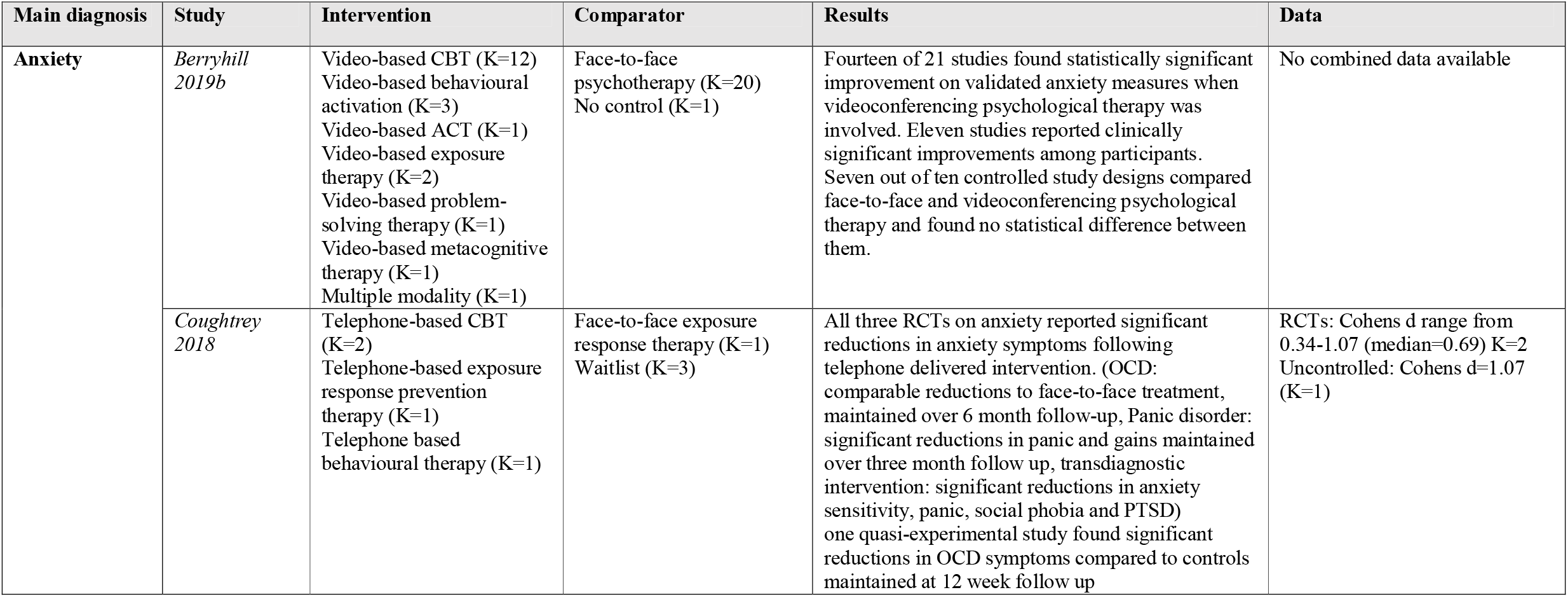
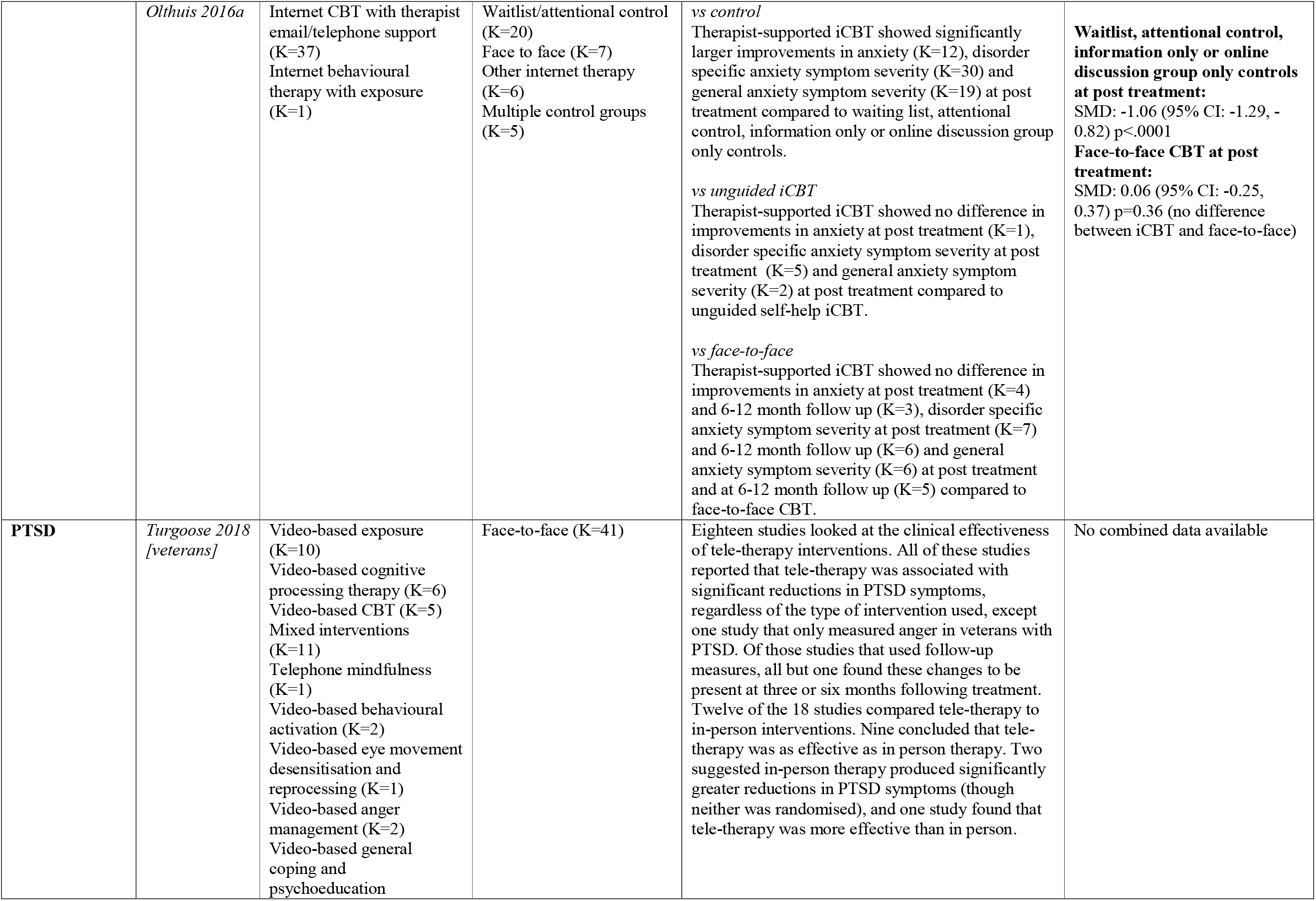
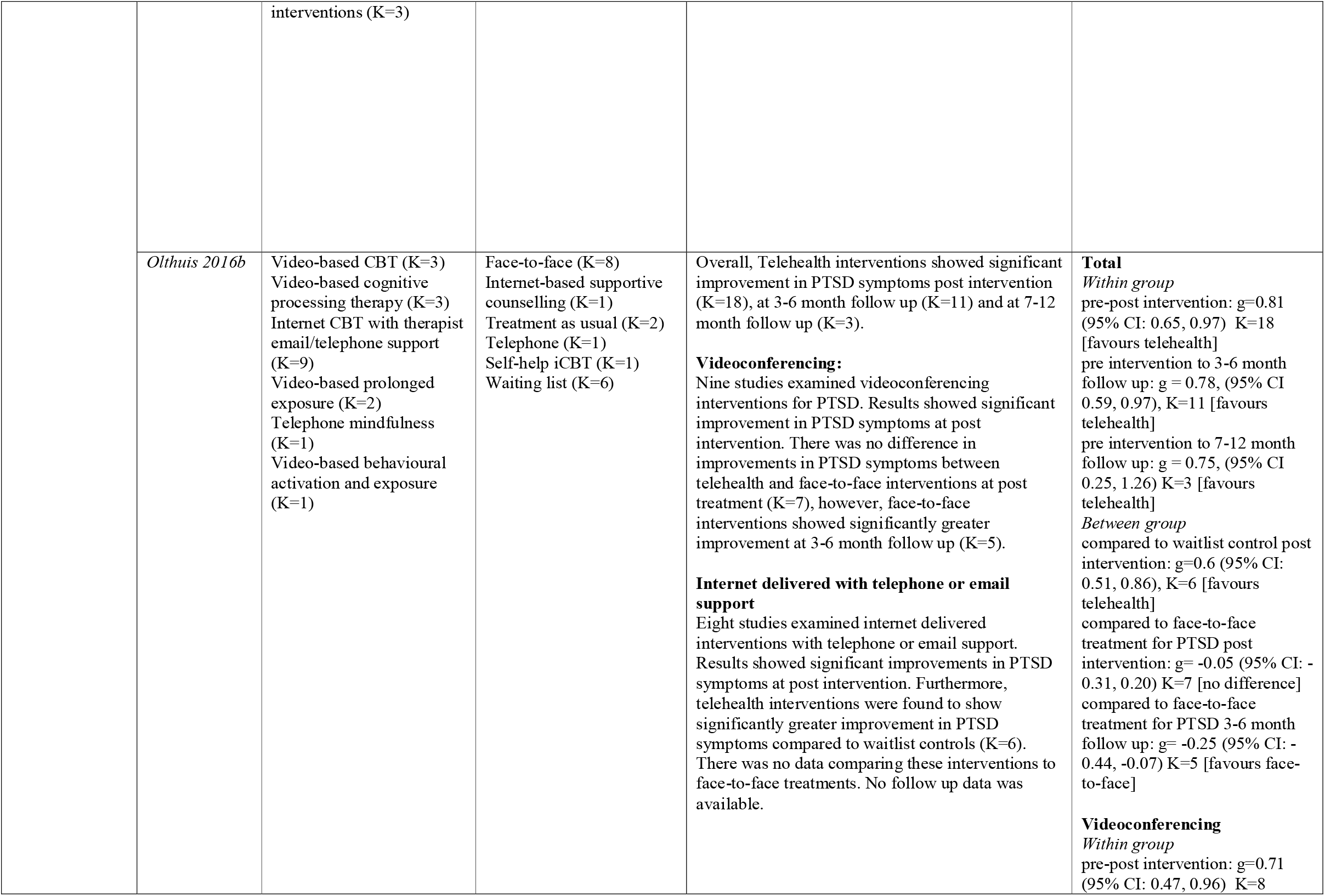
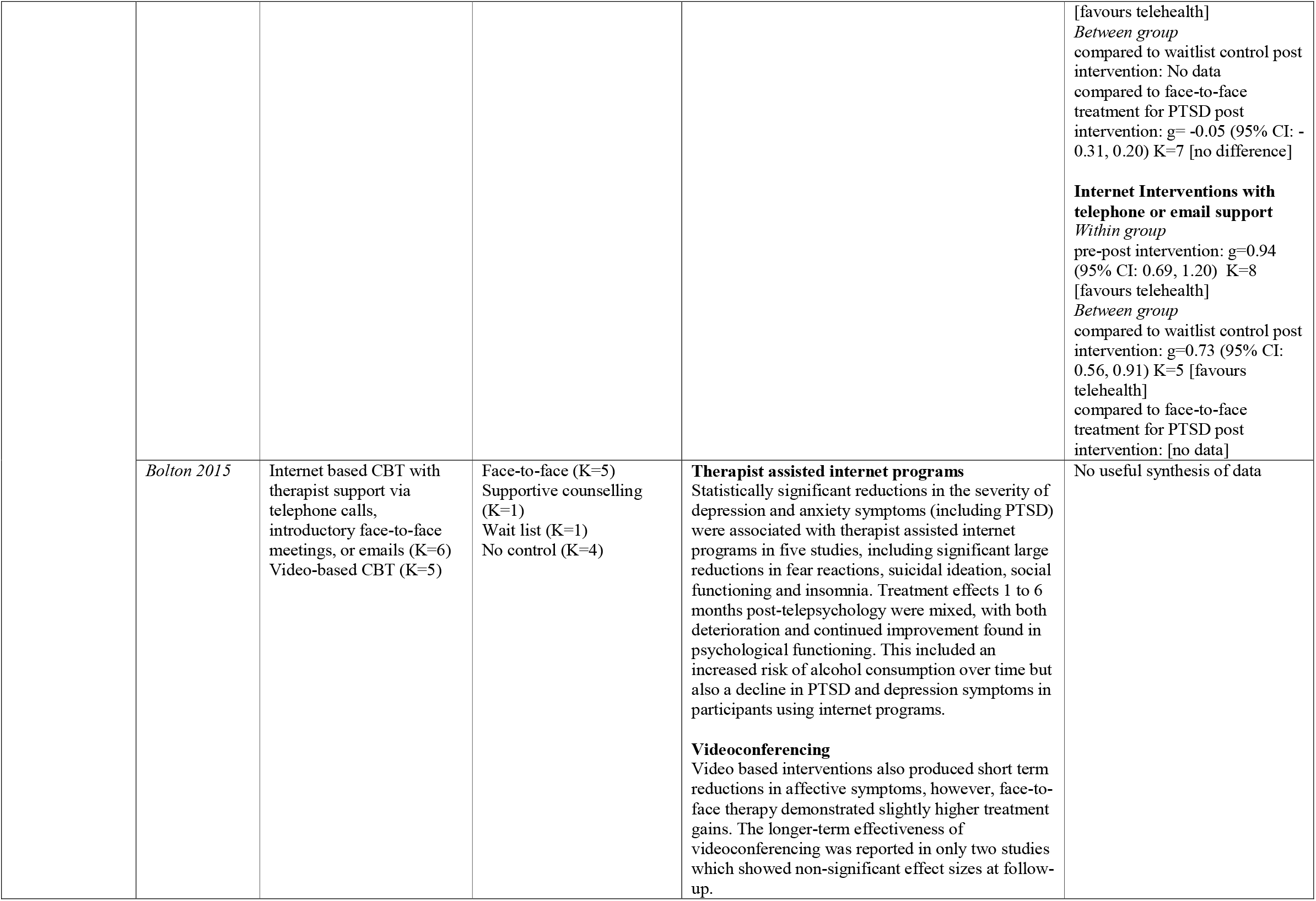
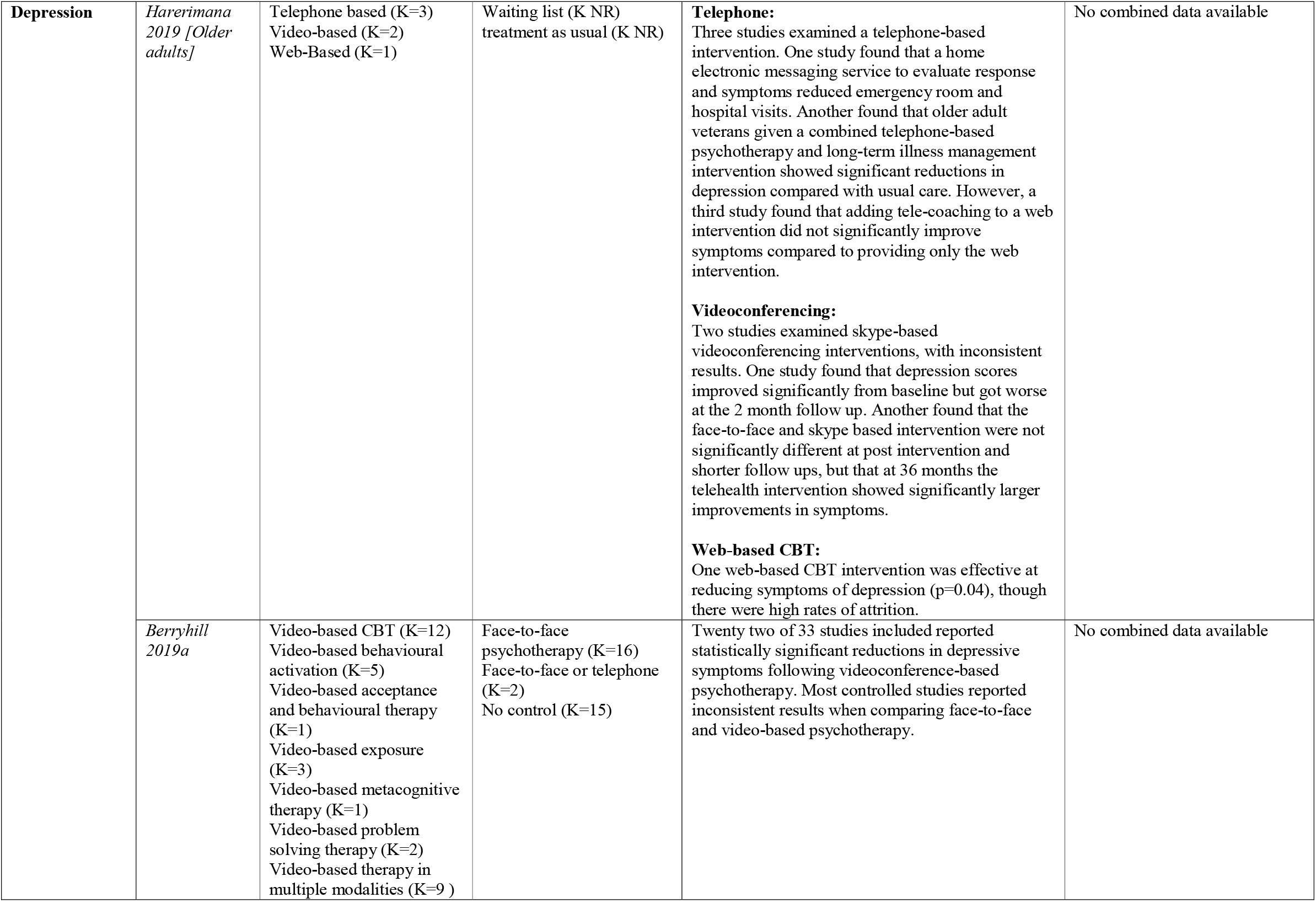
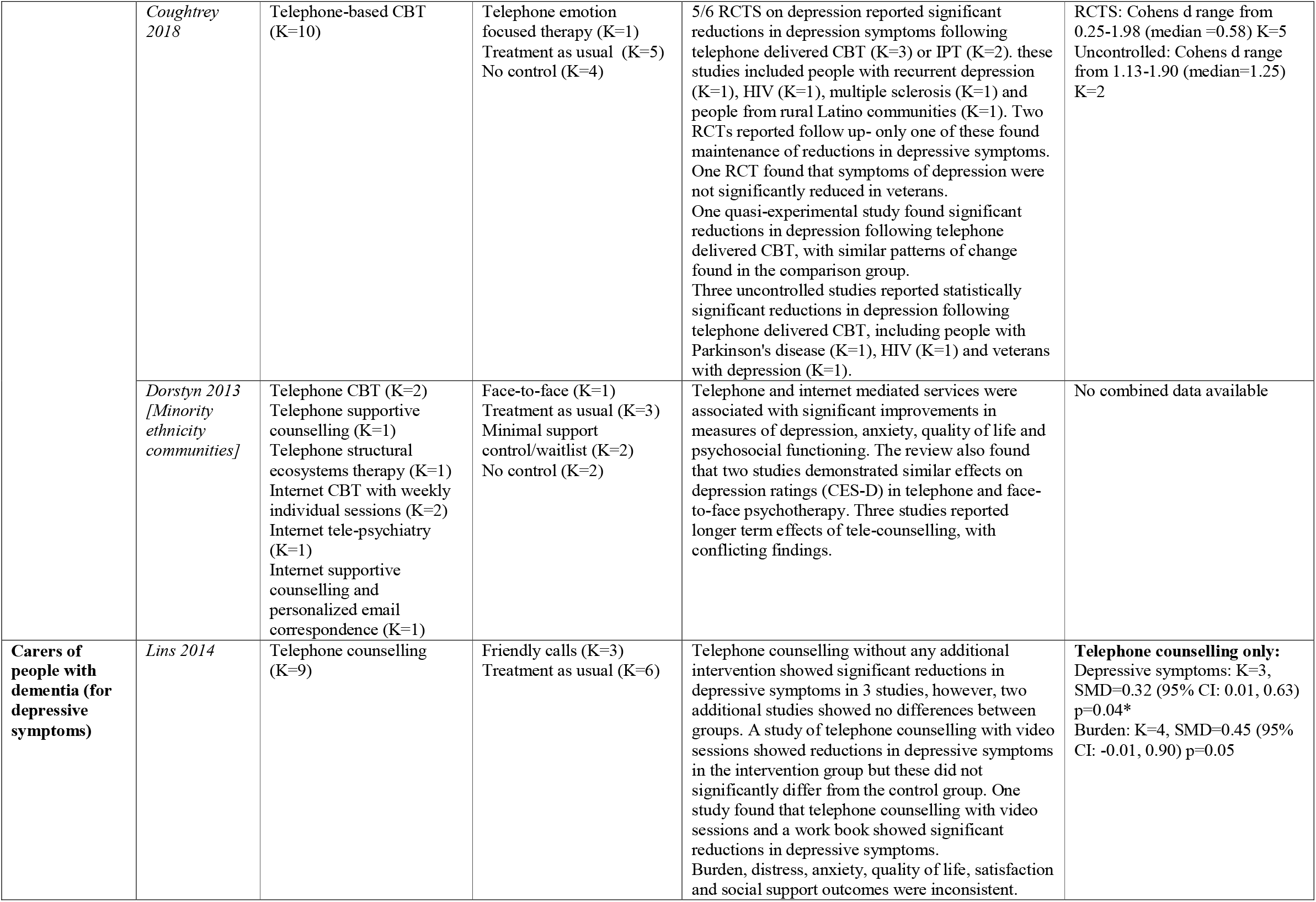
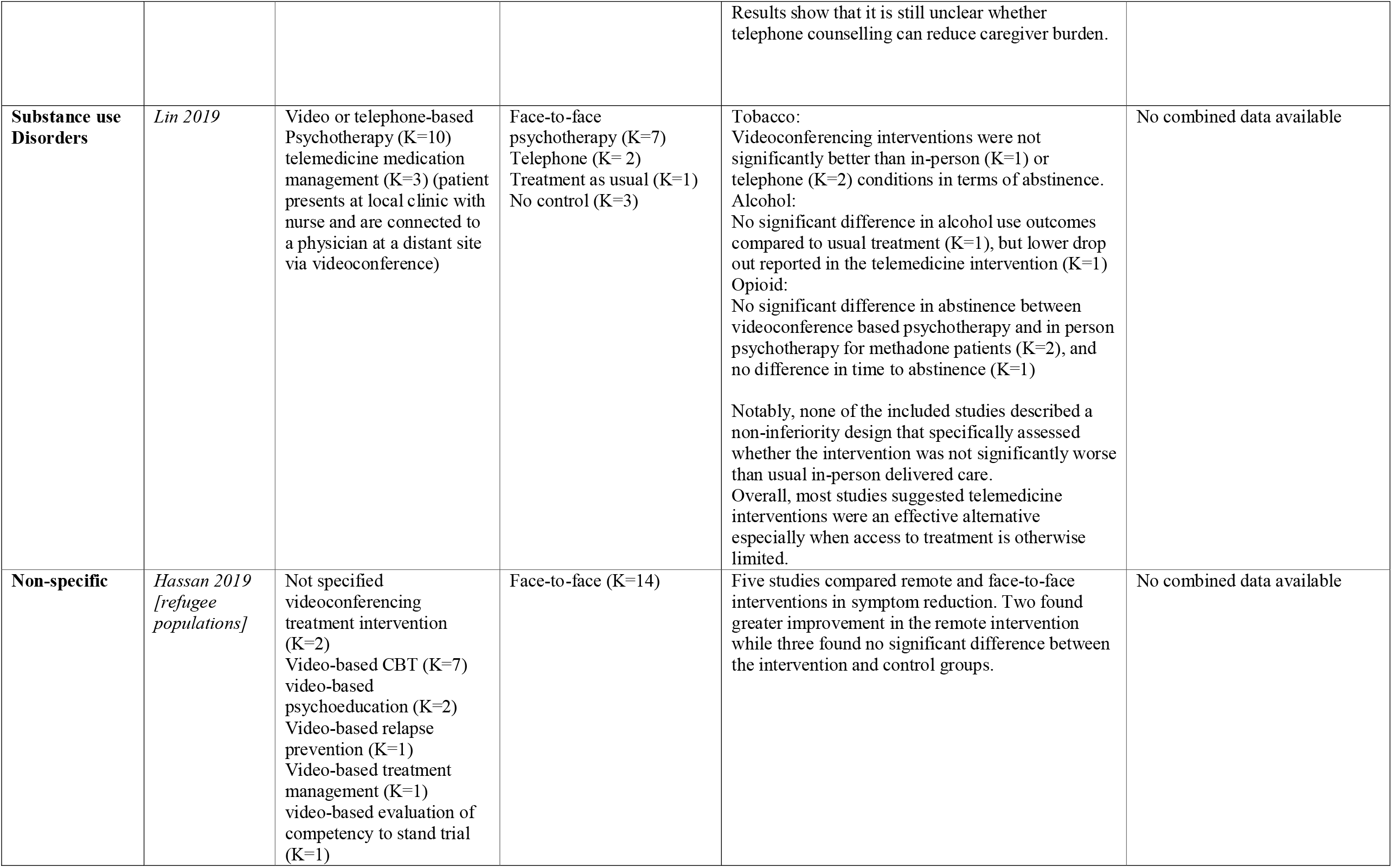
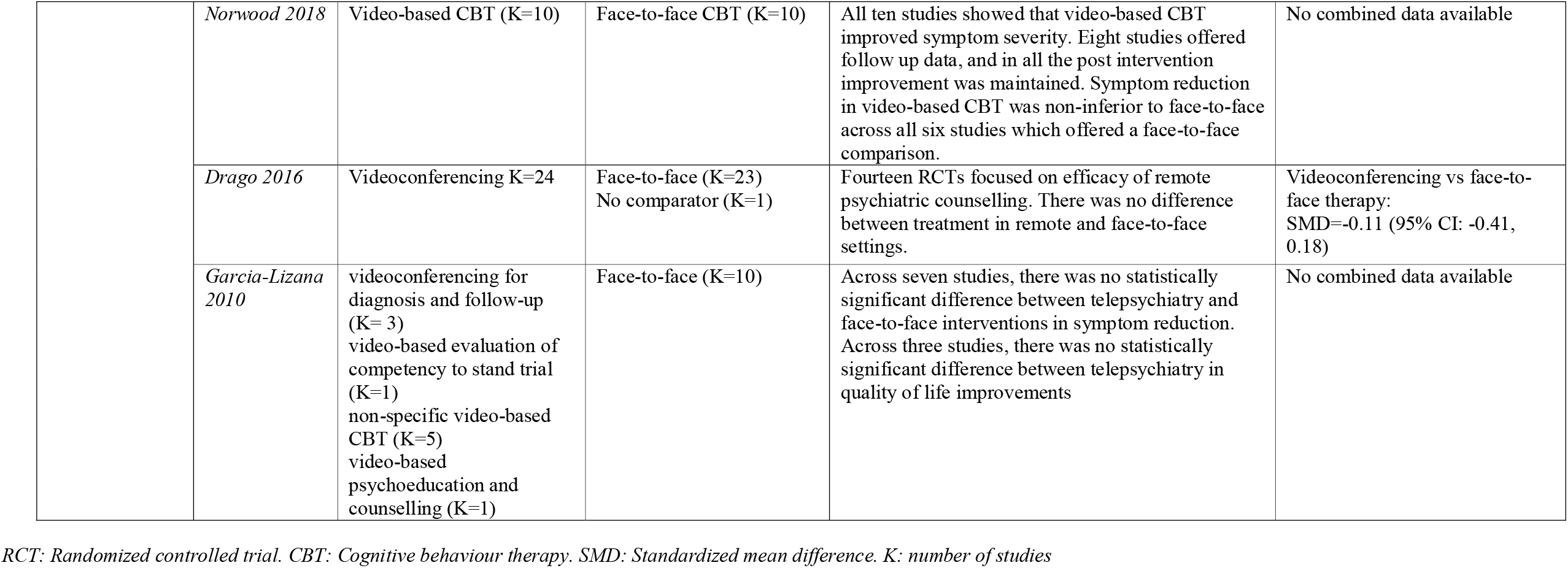
Clinical effectiveness outcomes.

### Implementation outcomes

Implementation outcomes were reported in 8 reviews (20, 22, 25, 26, 31, 33-35) Relevant outcomes included assessment comparability (K=2), fidelity to intervention and competence of therapists (K=1) (34), patient adherence to intervention (K=3) (20, 26, 31), patient attendance (K=4) (31, 33-35), safety (K=2) (26, 34), and technical difficulties (K=3) (26, 34, 35).

#### Assessment comparability

Limited evidence from one review suggests that video-conferencing can be used to provide assessment which is consistent with face-to-face assessment, with a correlation coefficient of 0.73 (95% CI: 0.63, 0.83) between conclusions of videoconference assessments and face to face assessments (25). A review of telephone assessments found that properly performed studies on telephone assessments were lacking, though telephone assessment for research purposes was suggested to have some potential use (22).

#### Fidelity and competence of therapists

One review (34) found that three studies of interventions for PTSD in veterans had been conducted that found fidelity and competence comparable to face-to-face interventions.

#### Patient adherence to intervention

Of three reviews (20, 26, 31) examining patients’ adherence to remote interventions, the general consensus was that comprehension of tasks and completion rates are high during both telephone and video-based CBT. However, one review found mixed findings, with one of the two studies it included reporting better adherence in the face-to-face intervention group for patients with PTSD, but equivalent adherence in remote and face-to-face conditions was found in another study of patients with depression. (20)

#### Patient attendance

Increased uptake and access to care compared to before use of remote technology was reported in reviews of depression treatment in older adults (18), PTSD treatment in veterans (34), and substance use disorder treatment (33). Drop out tended to be comparable to face-to-face interventions (33, 34). However, one review included a study reporting difficulty reaching ethnic minority patients with depression (31)

#### Safety

Patient safety when using remote interventions was reported in reviews of PTSD populations only. Two reviews agreed that safety was acceptable, with one reporting that generally with correct steps taken, safety could be managed in remote settings (34), and another reporting that client safety was deemed satisfactory (however no further detail was provided on this) (26).

#### Technical difficulties

Three reviews reported technical difficulties, none of which were identified as severe barriers to remote technology implementation. A review of older adults with depression found that four studies reported mistrust in technology (35), while more logistical challenges such as low image resolution and connectivity problems were reported in a review of video-based PTSD intervention for veterans (34). Another review reported findings from one included study that participants preferred mobile apps to supplement remotely delivered support (26). Further details on implementation outcomes are available in Table 3.

**Table 3:** Implementation outcomes.

| Outcome | Study | Assessment/Treatment | Main diagnosis | Intervention | Comparator | Results |
| --- | --- | --- | --- | --- | --- | --- |
| **Assessment comparability** | *Drago 2016* | Assessment and treatment | Multiple | Videoconferencing<br>K=24 | Face-to-face (K=23)<br>No comparator (K=1) | Assessment was found to be highly consistent between remote and face-to-face settings. Correlation coefficient=0.73 (95% CI: 0.63, 0.83) |
|  | *Muskens 2014* | Assessment | Multiple | Telephone diagnostic interviewing (K=16) | Face-to-face diagnostic interviewing (K=16) | There were too few studies which were properly performed to draw conclusions regarding the comparability of telephone and face-to-face interviews for psychiatric morbidity. Telephone interviewing for research purposes in depression and anxiety may however be a proper and valid method. |
| **Fidelity and competence of therapists** | *Turgoose 2018 [veterans]* | Treatment | PTSD | Video-based exposure (K=10)<br>Video-based cognitive processing therapy (K=6)<br>Video-based CBT (K=5)<br>Mixed interventions (K=11)<br>Telephone mindfulness (K=1)<br>Video-based behavioural activation (K=2)<br>Video-based eye movement desensitisation and reprocessing (K=1)<br>Video-based anger management (K=2)<br>Video-based general coping and psychoeducation interventions (K=3) | Face-to-face (K=41) | High levels of fidelity and therapist competence (K=3), with no significant differences compared to face-to-face. |
| **Patient adherence to intervention** | *Bolton 2015* | Treatment | PTSD | Internet based CBT with therapist support via telephone calls, introductory face-to-face meetings, or emails (K=6)<br>Video-based CBT (K=5) | Face-to-face (K=5)<br>Supportive counselling (K=1)<br>Wait list (K=1)<br>No control (K=4) | Qualitative feedback revealed that comprehension of the therapy materials was high, with participants completing set homework tasks (K=5) |
|  | *Dorstyn 2013*<br>*[Ethnic minorities]* | Treatment | Depression | Telephone CBT (K=2)<br>Telephone supportive counselling (K=1)<br>Telephone structural ecosystems therapy (K=1)<br>Internet CBT with weekly individual sessions (K=2)<br>Internet tele-psychiatry (K=1)<br>Internet supportive counselling and personalized email correspondence (K=1) | Face-to-face (K=1)<br>Treatment as usual (K=3)<br>Minimal support control/waitlist (K=2)<br>No control (K=2) | Most studies reported good treatment adherence with rates of completion of 75-97% |
|  | *Garcia-Lizana 2010* | Assessment and treatment | Multiple | videoconferencing for diagnosis and follow-up (K= 3)<br>video-based evaluation of competency to stand trial (K=1)<br>non-specific video-based CBT (K=5)<br>video-based psychoeducation and counselling (K=1) | Face-to-face (K=10) | Across two studies, mixed results were found for treatment adherence, with one study finding no difference and another reporting better adherence in the face-to-face group. |
| **Patient Attendance** | *Dorstyn 2013*<br>*[Ethnic minorities]* | Treatment | Depression | Telephone CBT (K=2)<br>Telephone supportive counselling (K=1)<br>Telephone structural ecosystems therapy (K=1)<br>Internet CBT with weekly individual sessions (K=2)<br>Internet tele-psychiatry (K=1) | Face-to-face (K=1)<br>Treatment as usual (K=3)<br>Minimal support control/waitlist (K=2)<br>No control (K=2) | One study reported difficulty reaching participants by telephone resulting in fewer sessions completed |
|  |  |  |  | Internet supportive counselling and personalized email correspondence (K=1) |  |  |
|  | *Christensen 2019 [Older adults]* | Treatment | Depression/Range of diagnoses including depression | Video consultations for tele-psychiatry (K=21) | F2F (11), no control (10) | Video consultations increased access to care and removed barriers such as having to travel (K=4). |
|  | *Lin 2019* | Treatment | Substance use Disorders | Video or telephone-based Psychotherapy (K=10)<br>telemedicine medication management (K=3)<br>(patient presents at local clinic with nurse and are connected to a physician at a distant site via videoconference) | Face-to-face psychotherapy (K=7)<br>Telephone (K= 2)<br>Treatment as usual (K=1)<br>No control (K=3) | Most studies reported increased retention in telemedicine groups (K=4) however no difference in in number of sessions attended was sometimes reported (K=2)<br>One alcohol study reported lower drop out in the telemedicine group, and more patients in this group were still in treatment at 6 months and one year. Two Opioid studies found that videoconference interventions resulted in better retention of participants up to one year compared to those receiving in person care. Another opioid study found >50% retention at 12 weeks but did not have a comparison group. However, another two studies found no difference between videoconference delivered psychotherapy and in person psychotherapy in the number of sessions attended |
|  | *Turgoose 2018 [veterans]* | Treatment | PTSD | Video-based exposure (K=10)<br>Video-based cognitive processing therapy (K=6)<br>Video-based CBT (K=5)<br>Mixed interventions (K=11)<br>Telephone mindfulness (K=1)<br>Video-based behavioural activation (K=2)<br>Video-based eye movement<br>desensitisation and<br>reprocessing (K=1)<br>Video-based anger<br>management (K=2)<br>Video-based general<br>coping and<br>psychoeducation<br>interventions (K=3) | Face-to-face (K=41) | In the majority of cases there were no differences between tele-therapy and in-person treatments on drop out or attendance. There was some indication that tele-therapy may help to increase uptake. |
| **Safety** | *Bolton 2015* | Treatment | PTSD | Internet based CBT with<br>therapist support via<br>telephone calls,<br>introductory face-to-face<br>meetings, or emails<br>(K=6)<br>Video-based CBT (K=5) | Face-to-face (K=5)<br>Supportive<br>counselling (K=1)<br>Wait list (K=1)<br>No control (K=4) | Client safety was deemed satisfactory |
|  | *Turgoose*<br>*2018*<br>*[veterans]* | Treatment | PTSD | Video-based exposure<br>(K=10)<br>Video-based cognitive<br>processing therapy<br>(K=6)<br>Video-based CBT (K=5)<br>Mixed interventions<br>(K=11)<br>Telephone mindfulness<br>(K=1)<br>Video-based behavioural<br>activation (K=2)<br>Video-based eye<br>movement<br>desensitisation and<br>reprocessing (K=1)<br>Video-based anger<br>management (K=2)<br>Video-based general<br>coping and<br>psychoeducation<br>interventions (K=3) | Face-to-face (K=41) | There might be some occasions where veterans have concerns about exposure tasks due to the lack of physical presence of the therapist, however overall it was established that these can be used just as effectively remotely. If appropriate steps are taken to manage safety, episodes of acute suicidality can also be managed. |
| **Technical difficulties** | *Bolton 2015* | Treatment | PTSD | Internet based CBT with therapist support via telephone calls, introductory face-to-face meetings, or emails (K=6)<br>Video-based CBT (K=5) | Face-to-face (K=5)<br>Supportive counselling (K=1)<br>Wait list (K=1)<br>No control (K=4) | Minimal technical difficulties were encountered (K=1)<br>participants reported that they would have preferred different forms of media, for example a mobile application, to supplement support (K=1) |
|  | *Christensen 2019 [Older adults]* | Treatment | Depression/Range of diagnoses including depression | Video consultations for tele-psychiatry (K=21) | F2F (11), no control (10) | Challenges such as mistrust in technology were reported frequently (K=4) |
|  | *Turgoose 2018 [veterans]* | Treatment | PTSD | Video-based exposure (K=10)<br>Video-based cognitive processing therapy (K=6)<br>Video-based CBT (K=5)<br>Mixed interventions (K=11)<br>Telephone mindfulness (K=1)<br>Video-based behavioural activation (K=2)<br>Video-based eye movement desensitisation and reprocessing (K=1)<br>Video-based anger management (K=2)<br>Video-based general coping and psychoeducation interventions (K=3) | Face-to-face (K=41) | Commonly reported technical difficulties were low image resolution on videoconferencing technology, not being able to connect, and audio delays. |

*RCT: Randomized controlled trial. CBT: Cognitive behaviour therapy. K: number of studies*

### Acceptability outcomes

Acceptability outcomes were reported in 10 reviews (18, 20, 21, 26, 27, 31-35). Relevant outcomes included clinician satisfaction (K=5) (18, 20, 21, 32, 34), therapeutic alliance (K=6) (21, 26, 27, 33-35), patient satisfaction (K=7) (20, 21, 31-35) and convenience (K=3) (21, 33, 35).

#### Clinician satisfaction

Overall, clinicians tend to report a preference for face-to-face interventions for both assessment and treatment (20, 32). However, some reviews have reported that clinicians find video-based therapies acceptable (32, 34). One review of remote interventions for carers of people with dementia found that counsellors felt they might need more support via debriefing following remote counselling sessions, and they also reported problems when reactions of carers could not be ascertained via the remote technology, and feelings of helplessness due to the impersonal nature of remote technology (21). Healthcare providers using remote interventions in older adults noticed practical benefits of telehealth (18).

#### Therapeutic alliance

Overall, good therapeutic alliance was reported as comparable to face-to-face interventions. However, some patient groups were found to feel more comfortable talking to therapists face-to-face, if possible, such as female older adults (35) or veterans (34). Meta-analysis was conducted in one review, which found that while standardized mean differences in alliance ratings were not significantly different, the lower limit of the 95% CI fell outside the pre-specified limit of non-inferiority, indicating that videoconference interventions may be inferior to face to face treatment, likely the result of therapist rated (but not patient rated) alliance scores being lower in the videoconference groups (27).

#### Patient satisfaction

High patient satisfaction was generally reported across seven reviews and patients tended to find remote interventions as satisfactory as face-to-face alternatives. This was true in substance use disorder (33), depression (20, 31, 32, 35), PTSD (34), older adult (35), ethnic minority (31), and carers of dementia patient populations (21), although Hassan et al. (32) reported a minority of studies indicating preference for face-to-face interventions. A review in older people noted that initial scepticism among both service users and providers tended to dissipate following positive experiences of video-conferencing, and that, with appropriate support and access to technology, even some who had not previously used computers reported positive experiences of video-calls (35). Accepting the need for treatment to be in tele-therapy form instead of face-to-face was reported as important in a study of veterans with PTSD (34)

#### Convenience

Patients reported the benefits of added convenience of therapy sessions at home via remote interventions for both depression (21, 35) and substance use disorders (33). Further details on acceptability outcomes are available in Table 4.

**Table 4:**
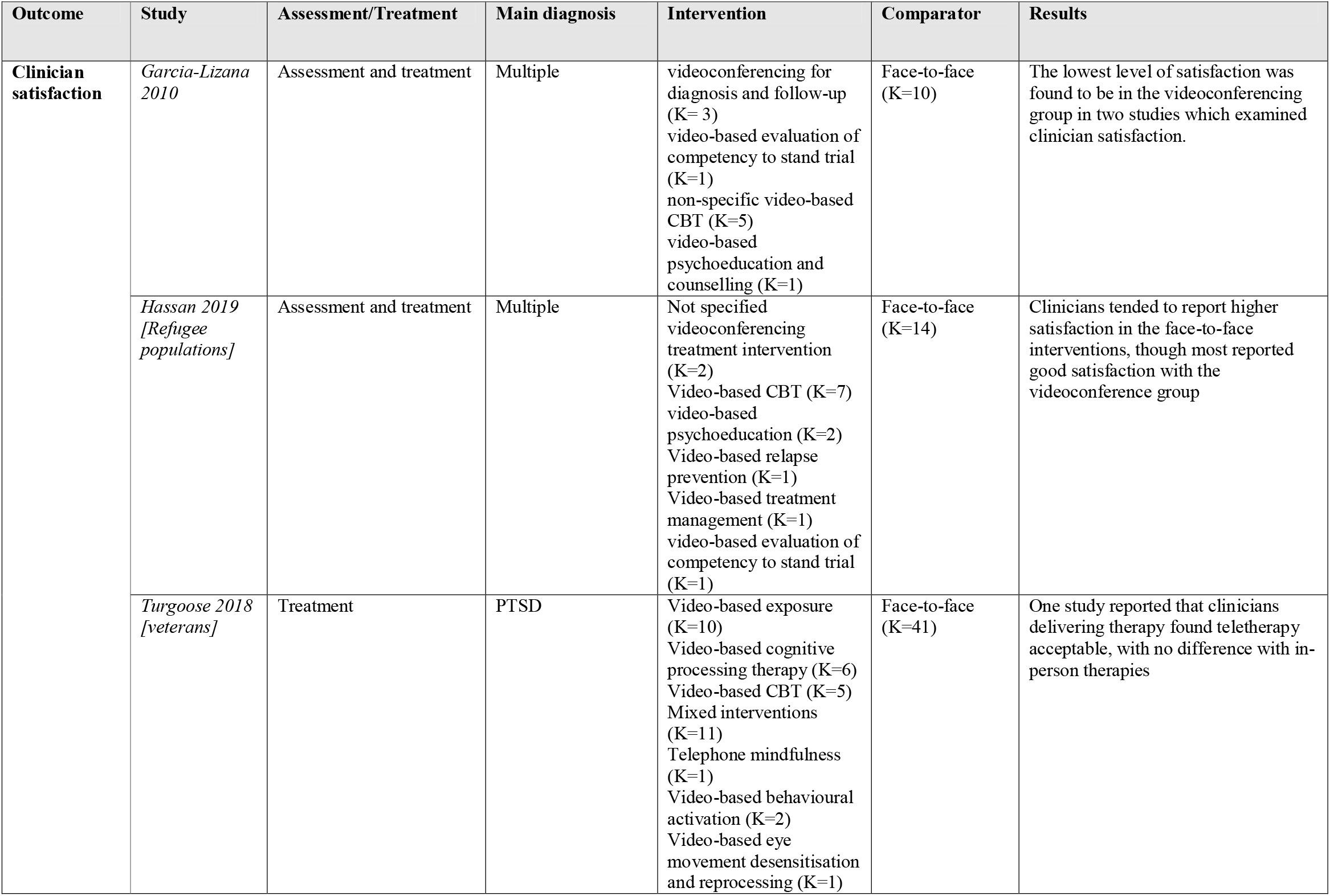
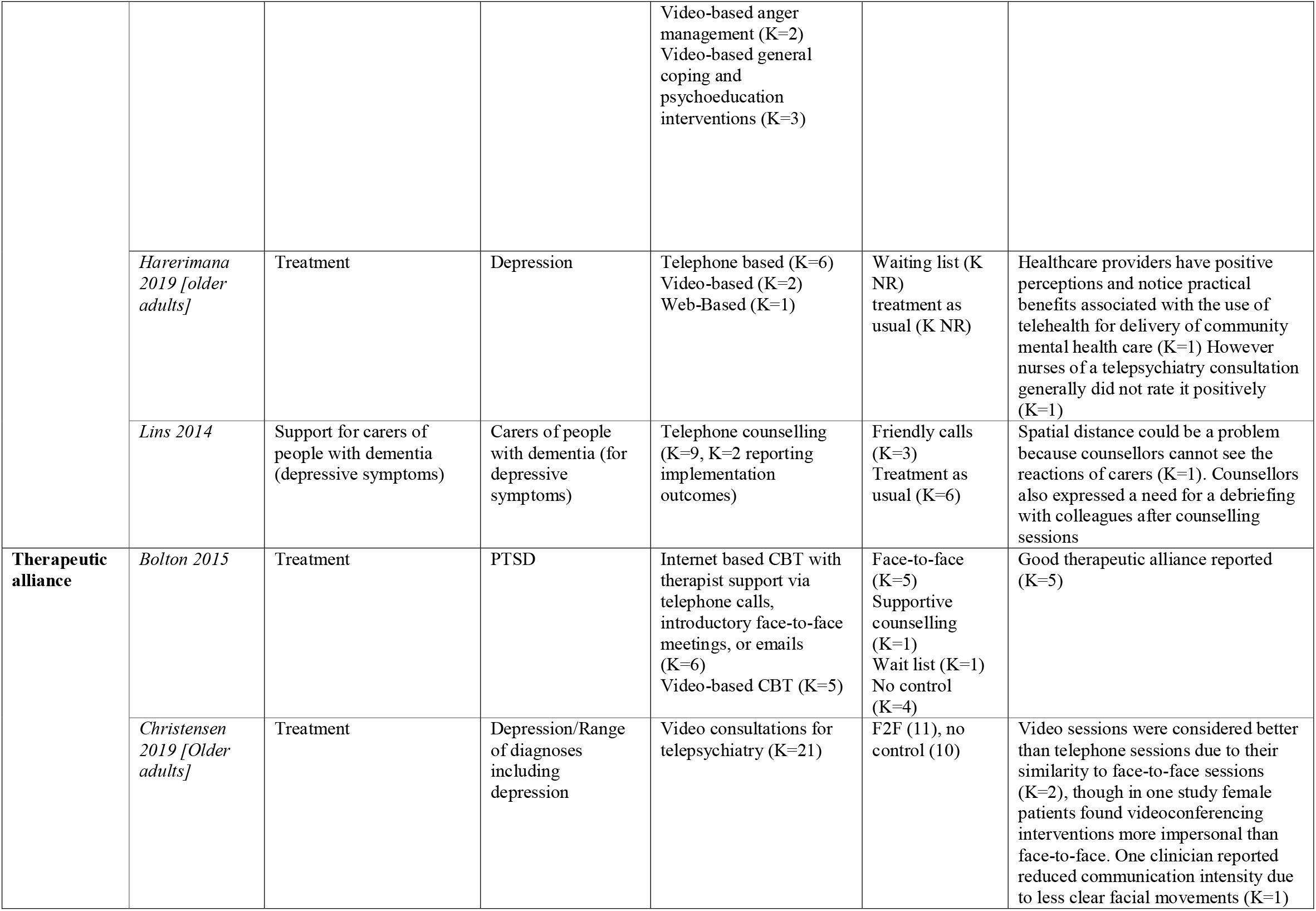
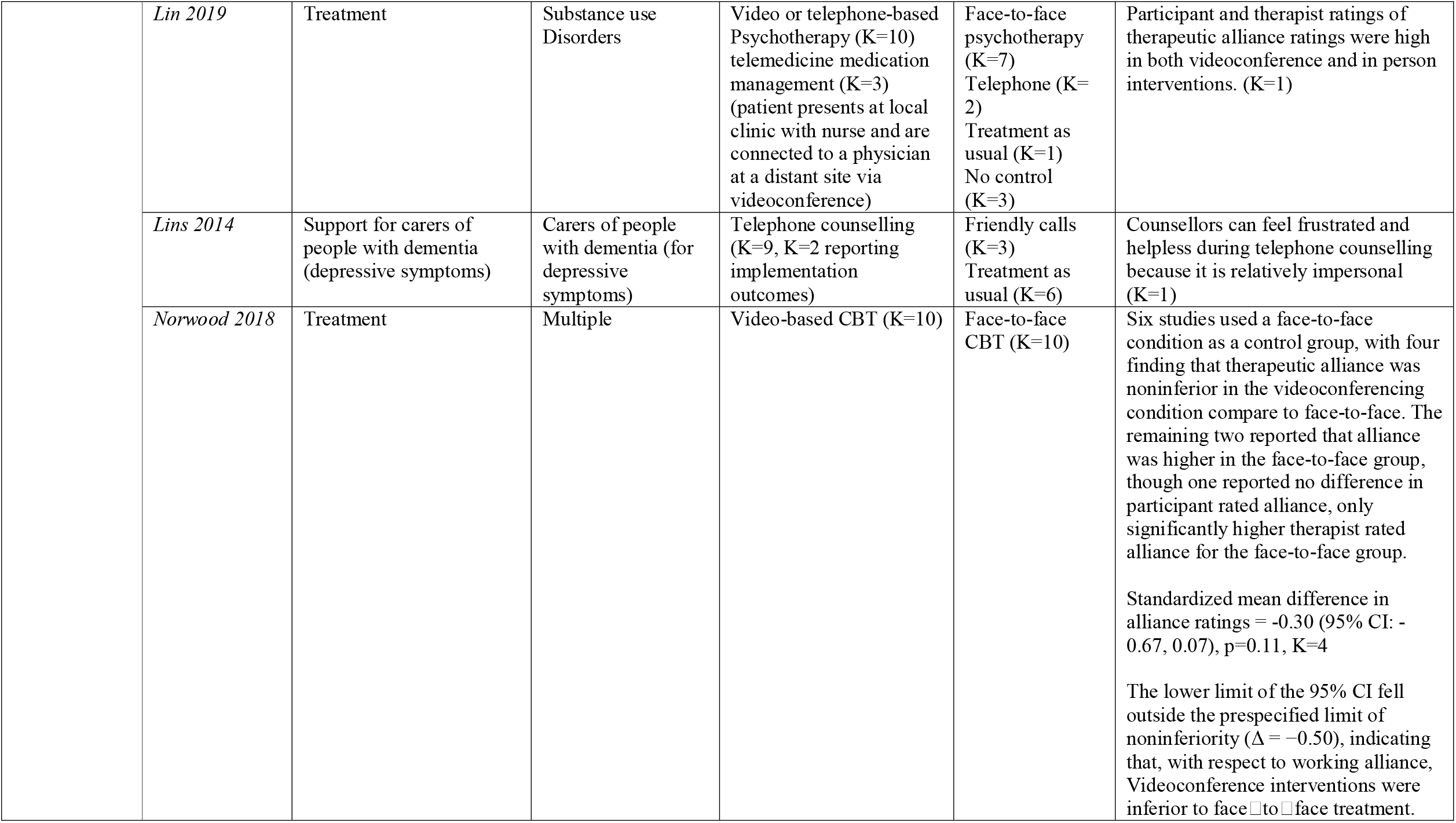
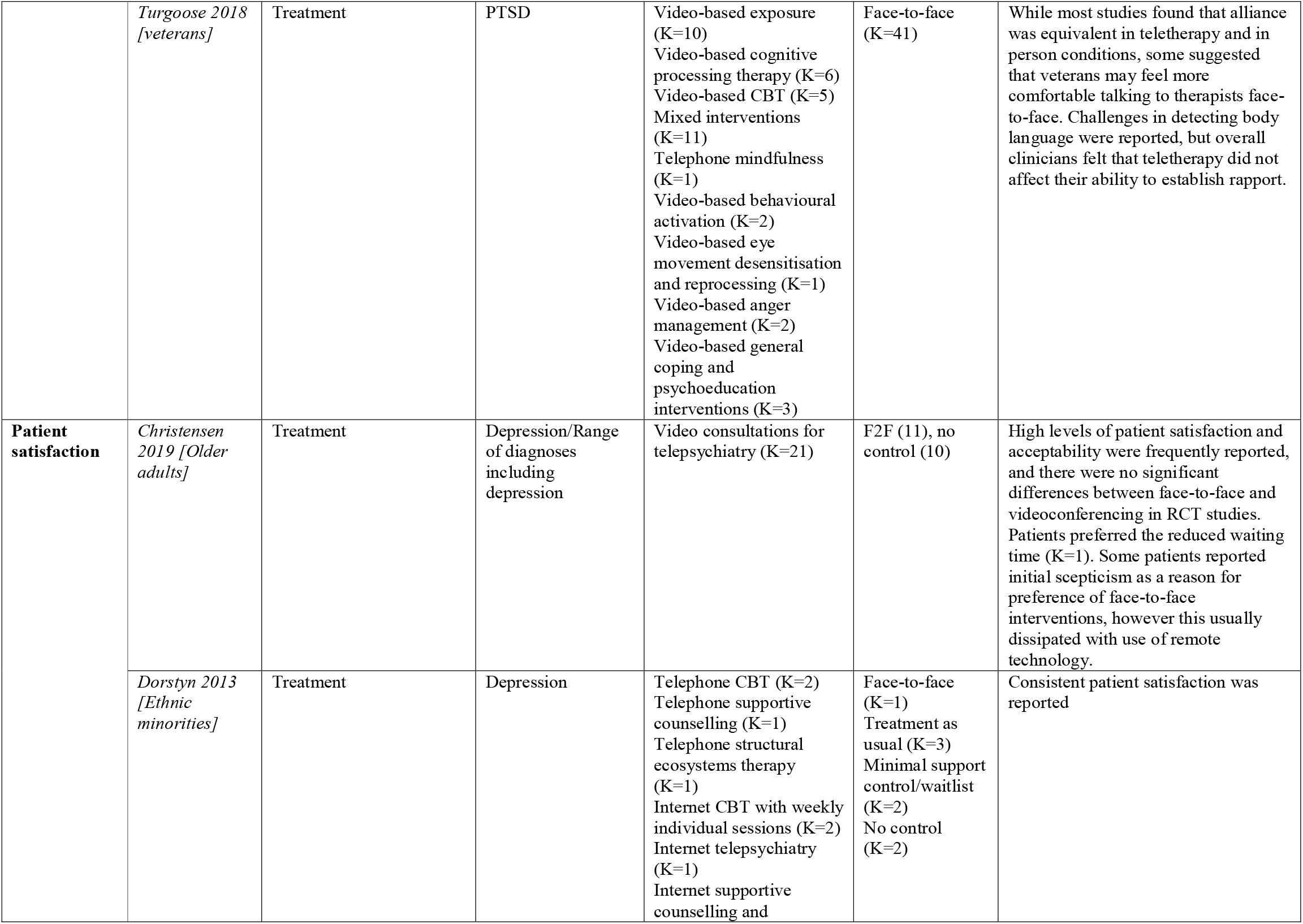
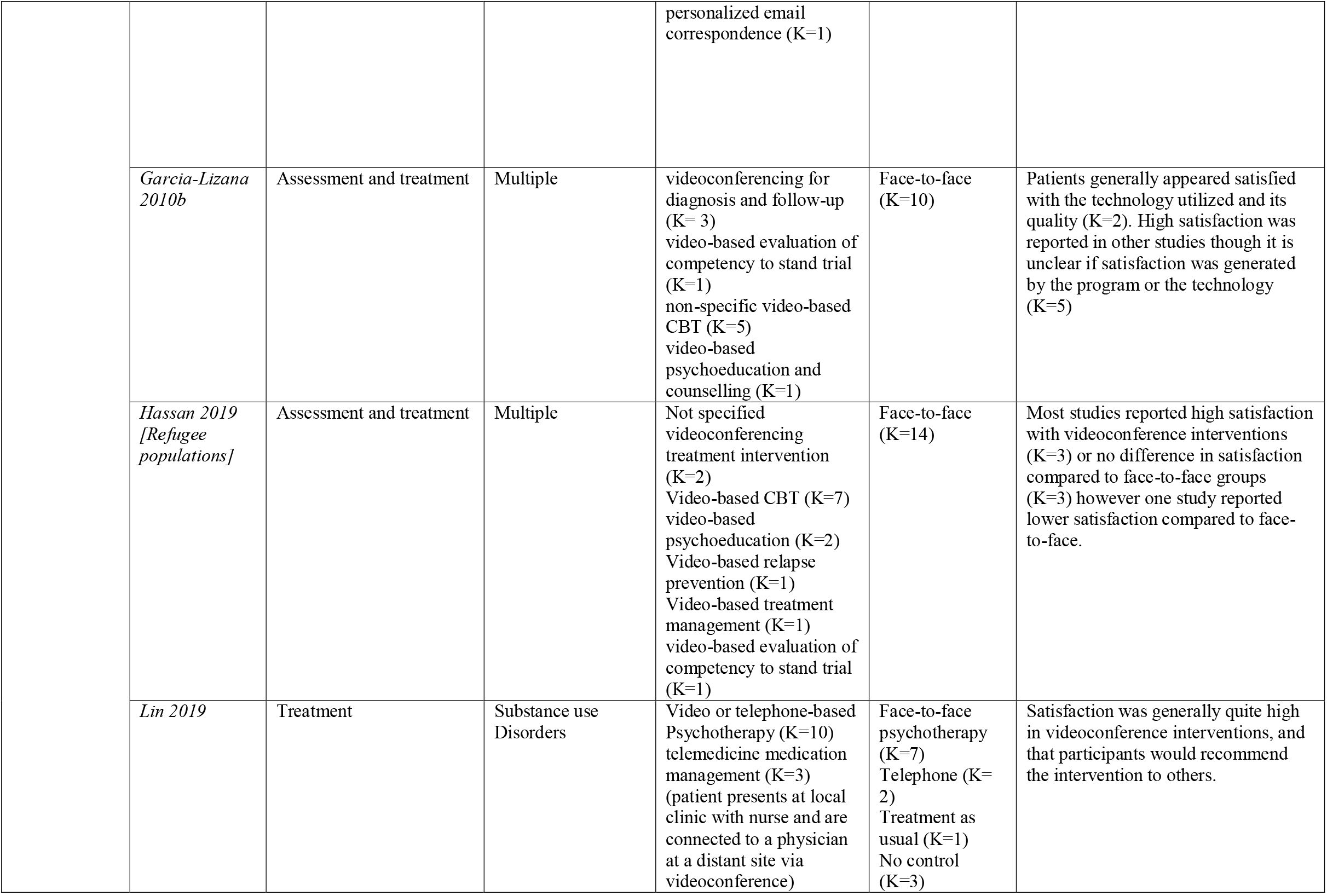
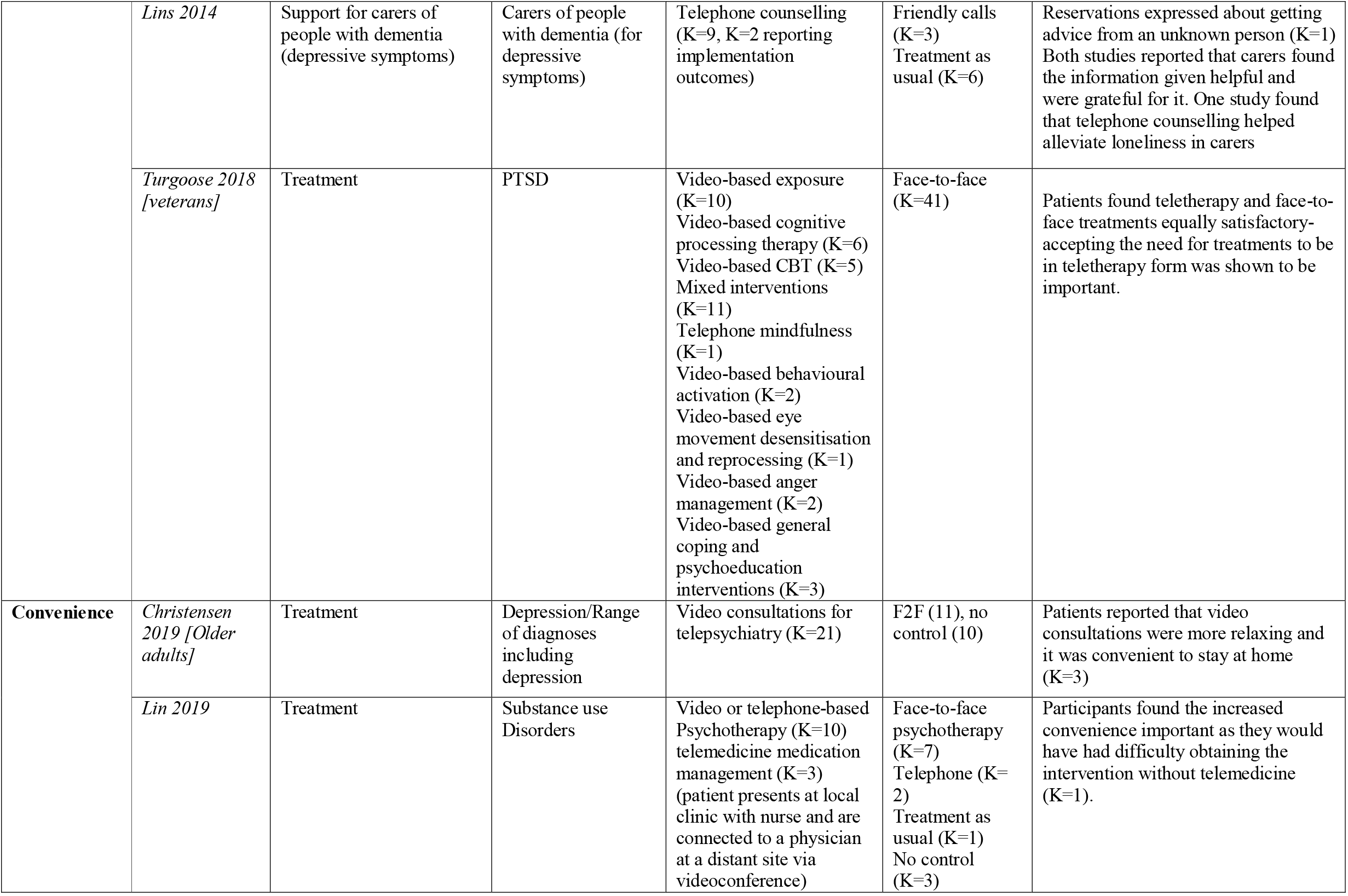
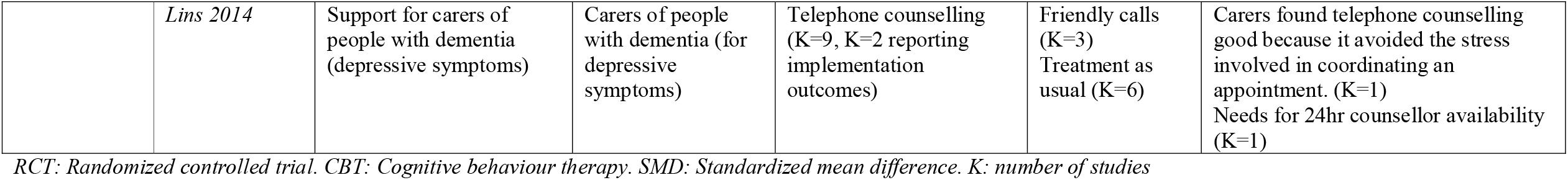
Acceptability outcomes.

### Cost effectiveness

Two reviews presented conclusions regarding the economic impact of telepsychiatry (32, 36). One review concluded that tele-psychiatry can be cost effective, compared to face-to-face interventions, particularly in rural areas where the number of consultations required before telepsychiatry becomes more cost effective (combatting initial equipment costs) is lower (32). The second review, whose main focus was on the cost effectiveness of telepsychiatry, reported that 60% (K=15) of included studies reported that telepsychiatry programmes were less expensive than standard in person care, due to savings such as travel time and reduced need for patients and their families to take time off work. However, eight studies concluded that telepsychiatry programmes were more expensive, particularly due to videoconferencing equipment costs. A final study included in the review found no difference in costs. The review also found a large range in reported costs, with, for example longer term delivery of telepsychiatry for Veterans ranging from $930 (2019 US dollars) to $2116 per patient. Cost effectiveness analyses were found in three included studies (37-39), which seemed to suggest that telepsychiatry was less cost effective. The review concluded that variation was due to large disparity in reporting of costs, for example whether personnel costs or initial equipment costs were included, and that there remains a need for future efforts to determine the cost effectiveness of different forms of telepsychiatry particularly for different disorders and applications of remote technology (e.g. consultation vs therapy). In addition, Dorstyn and colleagues (31) looked at health service utilisation which can impact cost effectiveness. They found that rates of antidepressant and health service utilisation were similar in the 3 months following both telephone and web-based counselling.

### Guidelines

Only one review (23) of guidelines for remote working was found that met the inclusion criteria. This review comprehensively summarised the guidance published to date, including guidance on decisions about the appropriateness of e-mental health, ensuring competence of mental health professionals, legal and regulatory issues, confidentiality, professional boundaries, and crisis intervention. Recommendations from 19 guidelines were characterized as either firm (50% or more recommending) or tentative (fewer than 50% recommending). The review identified as firm recommendations ensuring that remote interventions were appropriate for the needs of individual patients and within the boundaries of therapist competence, laws and regulations; maintaining confidentiality and seeking informed consent, including for specific aspects of remote appointments such as data security; and ensuring geographically accessible in-person clinical support is available in case of crisis or emergency. Guidelines suggested a higher risk of harm for people with cognitive impairments and psychotic disorders, but did not provide concrete recommendations as to how to adapt to these populations. Furthermore, a minority of guidelines discussed remote technology in young people, with the main message being the importance of checking consent with both the patient and parent. A full summary of recommendations from the review can be found in Appendix 4.

## Discussion

Our umbrella review retrieved a variety of recent relevant systematic reviews, on which future planning of tele-mental health implementation can usefully draw. Across the 19 reviews included in this umbrella review, results suggest that remote forms of assessment and intervention can produce at least moderate decreases in symptom severity for people suffering from a variety of mental health conditions. Arguments are strongest for videoconferencing interventions, with multiple reviews concluding that outcomes appear comparable to face-to-face interventions in the short term. However, at present, conclusions regarding longer term results remain uncertain: while some reviews have reported maintenance of positive effects at short term and long-term follow-ups for both videoconference and telephone-based interventions, other reviews have suggested that effects are less long-lasting than face to face intervention and the amount of evidence on which to base this assessment is limited

Reviews also suggest that remote interventions are satisfactory to service users participating in studies, who tended to report being as satisfied as with face-to-face interventions. This is promising in relation to adaptations during the COVID-19 crisis and for the future, but the reviews tend to relate to small-scale and carefully planned implementations of tele-mental health with volunteer participants, rather than to large-scale emergency implementations as in the current crisis. Clinician satisfaction varied more, with reviews tending to conclude that while remote interventions may be acceptable, face-to-face intervention is usually preferable. This may be related to reports in some reviews that clinician-ratings of therapeutic alliance are poorer with tele-mental health (27, 40). Despite this, patients tend to feel that alliance is on-par with face-to-face interventions (27, 33, 34). There is some suggestion that training and more experience with video and telephone-based technology for intervention delivery may alleviate this concern in therapists (40), although staff reports following increased uptake in the COVID crisis seem to suggest continued concerns about rapport (2).

Evidence yielded by reviews on the important questions of whether assessments appeared accurate and comprehensive and whether treatment was delivered as intended was limited. Two reviews examined comparability of remote versus face-to-face assessment, with one review finding good correlation between assessments, and another finding that there was insufficient high-quality evidence published thus far to draw accurate and meaningful conclusions (22, 25). Regarding fidelity, we found one review that reported good therapist fidelity and competence in remotely delivered interventions in the context of service delivery for veterans with PTSD (34): thus, there appears to be a gap in the evidence as reported in systematic reviews as to whether high fidelity and quality is achieved with tele-mental health interventions. High quality standardised training rooted in evidence will be important to ensuring high quality and overcoming self-doubt among clinicians in delivering remote interventions (23, 40, 41).

A crucial question regarding the rapid adoption of remote technologies during the pandemic has been how far service users may drop out of or be excluded from care as a result. A minority of the reviews included relevant data, most of it relatively reassuring. Reviews reported that remote interventions were convenient, and those examining uptake reported an increase. Where examined, retention was also comparable to face-to-face treatment (33, 34). Reports of technological difficulties were reassuringly few across reviews, although this may be more easily achieved with the well-planned, smaller-scale implementations of tele-mental health that characterise research studies than with larger scale implementation. However, one aspect of remote delivery in which reviews did not generally report is the risk of complete digital exclusion for those patients who may not have the skills or resources to engage with remote therapy or assessments (1, 2). Implementation of tele-mental health across service systems is only likely to be beneficial if there are clear plans for preventing patients with limited access to technology from being at a disadvantage (42, 43), whether by supporting them to engage with remote care or ensuring that equivalent care is available face-to-face.

Digital exclusion may result in the exacerbation of existing inequalities where already disadvantaged groups, such as older adults, people with sensory or cognitive impairment or members of some Black Asian and Minority Ethnic Groups, are at greater risk of exclusion (1, 44, 45). Some included reviews have examined this (18, 35). A single review by Dorstyn and colleagues (31) reported that members of predominantly North American ethnic minority communities with depression benefited from tele-counselling. To consolidate this further, a broader evidence base is thus urgently required to evaluate the risk of exacerbating ethnic inequalities in mental health care access through tele-mental health adoption. Furthermore, many have argued that the shift to remote working may exclude older adults (35, 44). With findings from one review (18) suggesting videoconferencing interventions can be comparable to face-to-face, and another (35) finding high levels of patient satisfaction, therapeutic alliance, attendance and convenience, this review suggests effective remote intervention delivery may be feasible for older adults. This is encouraging as staying at home and avoiding infection during the pandemic is especially desirable for older adults. No reviews were found regarding other sub-groups of potential concern, such as people with sensory or cognitive impairments, children and adolescents and their families or people with comorbid mental and physical health conditions. We also did not find substantial evidence on settings of particular interest, such as mental health inpatient services (including the use of tele-mental health in compulsory detention processes) and crisis services.

### Limitations

The findings of this umbrella review should be considered alongside a number of limitations. Firstly, umbrella reviews by their nature aim to present an overview of findings from systematic reviews (46), making conclusions reliant on the quality and reporting accuracy of included reviews and necessarily resulting in some loss of nuance when findings are pooled. Although we included only reviews considered to be systematic (defined here as searching at least three databases, and conducting a quality assessment when synthesising quantitative data), it was apparent from our quality assessment that the majority of reviews lacked several attributes characteristic of a high-quality review with robust conclusions, for example pre-specified protocols and duplicate study selection. However, our aim was to gain a rapid overview, relevant especially to current and future rapid implementation of tele-mental health, of the extent of supporting evidence to be drawn from previous literature regarding tele-mental health: the umbrella review provides a useful route to achieving this. Inclusion of systematic reviews focused on methods other than randomised controlled trials and on guidance further increases the methodological variability of included reviews and studies, but is a choice made to maximise retrieval of material from which real-world important lessons can be learnt regarding feasibility, acceptability and implementation barriers and facilitators (47).

This review also aimed to summarise outcomes relating to cost-effectiveness of remote delivery. We found only two reviews which summarised this outcome and only one which did this comprehensively. Given conclusions that further work should be done to establish the cost effectiveness of different forms of remote working, for different patient groups, there is a significant gap in the literature given that efficiency is one of the arguments made to support remote interventions (48).

Finally, this review aimed to summarise the literature published prior to the COVID-19 pandemic to identify evidence relevant both to the current context and the recovery from the pandemic. However, the current pandemic has given rise to a much more extensive switch to tele-mental health than previously, meaning that not all conclusions may be generalised to “the new normal”. In particular, the evidence retrieved in this review tends not to relate to implementation of tele-mental health across whole catchment areas and does not yield much evidence relevant to currently highly salient issues such as risks of digital exclusion or exacerbation of mental health inequalities and economic disadvantage which may well be exacerbated as a result of COVID-19 (1, 2). Conclusions of this review should be supplemented with further scrutiny of adoption of remote working within the context of these societal changes.

## Conclusion

Research across a range of mental health conditions suggests that tele-mental health is potentially an effective, feasible and acceptable tool for providing mental health treatment, at least when interventions are relatively well-designed and well-planned, as has tended to be the case in research studies. Comparability in terms of symptom improvement and satisfaction to face-to-face methods suggests the move to tele-mental health to sustain mental health services during the pandemic has probably been a reasonable one, although the context of this emergency implementation has been very different from most research studies. Further research should seek to build on existing evidence in establishing the longer-term effectiveness and cost-effectiveness of tele-mental health in a range of groups and settings, for example including children and young people and inpatient acute services and focusing on issues of inclusion and reach. A further question on which further evidence would be highly desirable is the extent to which digital exclusion can be remedied, including examination of interventions designed to include those with limited previous digital resources or skills. Future planning for tele-mental health implementation should draw both on previous research evidence, often acquired in relatively small-scale studies, and on COVID-19 learning from experiences of trying to engage large service user populations and most of the mental health workforce with remote technology delivery.

## Data Availability

Data extraction is available from the author on request.

