## Appendices for "Remote working in mental health services: a rapid umbrella review of pre-COVID-19 literature"

### Appendix 1: Search strategy

| **Psychinfo** | **604** |
| --- | --- |
| **CENTRAL systematic reviews** | **94** |
| **Pub med** | **543** |
| **Total** | **1241** |
| **Dupes removed** | **1086** |

**All searches 1^st^ January 2010-August 26^th^ 2020**

**PsychINFO**

[*reviews]*

1. (systematic or structured or evidence or trials or studies).ti. and ((review or overview or look or examination or update* or summary).ti. or review.pt.)
2. meta-analysis.pt. or (meta-analys* or meta analys* or metaanalys* or meta synth* or meta-synth* or metasynth*).ti,ab,id,hw.
3. ((systematic or meta) adj2 (analys* or review)).ti,id. or ((systematic* or quantitativ* or methodologic* or qualitative) adj2 (review* or overview*)).ti,ab,id,sh. or ((quantitativ* or qualitativ*) adj2 synthesis*).ti,ab,id,hw.
4. (integrative research review* or research integration).ti,ab. or (review.ti,id,pt. and (trials as topic or studies as topic).hw.) or (evidence adj3 review*).ti,ab,id. or (realist adj3 review*).ti,ab,id.
5. (OR/1-5)

[*mental disorders]*

6. mental disorders/ or anxiety disorders/ or obsessive compulsive disorder/ or panic disorder/ or phobias/ or social phobia/ or bipolar disorder/ or eating disorders/ or anorexia nervosa/ or binge eating disorder/ or bulimia/ or affective disorders/ or major depression/ or (exp personality disorders/) or schizophrenia/ or affective psychosis/ or catatonic schizophrenia / or paranoid schizophrenia/ or body dysmorphic disorder/ or posttraumatic stress disorder/ or delusions/ or dysthymic disorder/ or endogenous depression/ or reactive depression/ or recurrent depression/ or treatment resistant depression/ or atypical depression/ or “depression (emotion)”/ or self-injurious behavior/ or suicidal ideation/ or attempted suicide/
7. alzheimer's disease/ or exp dementia/
8. (affective disorder* or agoraphobi* or anorexia nervosa or anxiety or BPD or binge eat* or binging or bipolar or bulimi* or combat disorder* or compulsi* or delusion* or depersonali#ation or depressed or depression or depressive or eating disorder* or EDNOS or emotional trauma or mania or manic or mood? or neurotic or obsess* or panic or paranoi* or parasuicid* or personality disorder* or phobi* or ((post-trauma* or posttrauma*) adj stress*) or psychiatr* or psychopathol* or psychosomatic or psychotic or psychos* or PTSD or schizo* or (self adj (injur* or harm or mutilat*)) or social anxiety or suicid*).ti,id,hw.
9. Mental health/ or ((mental* or psychiatric) adj2 (health* or ill* or disorder* or diagnos?s or problem*)) .ti,ab,id.
10. Mental health services/ or community mental health services/ or community psychiatry/ or (mental health service? or CAMHS or "child and adolescent mental health service?" or psychiatry or psychology or psychotherap*).ti,ab,id.
11. (OR/6-10)

[*remote working]*

12. telecommunications/ or telepsychiatry/ or telemedicine/ or computer assisted therapy/ or telephone/ or technology/ or videoconferencing/ or internet/ or computer mediated communication/ or computers/ or exp online therapy/
13. *Answering service*.ti, id.*
14. ((mobile* or phone? or telephone? or remote* or distan* or online or virtual or electronic or email or e-mail or video) adj3 (consult* or counsel* or follow up or follow-up or support* or interview* or monitor* or therap* or treatment? or "CBT")).ti,ab,id.
15. (ehealth* or e-health* or emedicine* or e-medicine* or etherap* or e-therap* or mhealth or m-health or m health or eCBT or e-CBT or iCBT or i-CBT or Interap* or (electronic adj2 CBT) or telemedicine or telecare or telepsychiatry or telecommunications or teleconferencing or computer assisted therap* or teletherap* or telemental or tele-mental).ti,ab,id.
16. *(videoconferen* or video-conferen* or videophone? Or video-phone? or video-call* or video-based call* or video call* or video based call*).ti, ab, id.*
17. *Digital interventions/*
18. *Exp electronic communication/*
19. *Mobile devices/ or mobile health/ or mobile technology/*
20. *Teleconferencing/*
21. *(OR/12-20)*
22. *5 AND 11 AND 21*

*Restrict: 2010-2020*

**PUB MED**

**("systematic review"[Title/Abstract] OR "literature review"[Title/Abstract] OR "narrative review"[Title] OR "qualitative review"[Title] OR "evidence review"[Title] OR "systematic quantitative review"[Title] OR "meta review"[Title] OR "systematic critical review"[Title] OR "realist review"[Title] OR"systematic cochrane review"[Title] OR "systematic search and review"[Title] OR "systematic integrative review"[Title] or "qualitative synthesis"[Title] or "narrative synthesis"[Title] or meta-synthesis*[Title/Abstract]) AND (("Dementia"[MeSH Terms] OR "Alzheimer Disease"[MeSH Terms]) OR ("Mental Health"[Title/Abstract] OR "mental problem*"[Title/Abstract] OR "mental disorder*"[Title/Abstract] OR "mental illness*"[Title/Abstract] OR "Depression"[Title/Abstract] OR "depressive disorder*"[Title/Abstract] OR "Anxiety"[Title/Abstract] OR "anxiety disorder*"[Title/Abstract] OR "phobi*"[Title/Abstract] OR "agoraphobi*"[Title/Abstract] OR "anxious"[Title/Abstract] OR "obsess*"[Title/Abstract] OR "compulsi*"[Title/Abstract] OR "panic"[Title/Abstract] OR "PTSD"[Title/Abstract] OR "post traumatic stress"[Title/Abstract] OR "posttraumatic stress"[Title/Abstract] OR "stress disorder*"[Title/Abstract] OR "psychiatr*"[Title/Abstract] OR "SMI"[Title/Abstract] OR "psycho*"[Title/Abstract] OR "schizo*"[Title/Abstract] OR "manic"[Title/Abstract] OR "mania"[Title/Abstract] OR "bipolar"[Title/Abstract] OR "personality disorder*"[Title/Abstract] OR "self-harm"[Title/Abstract] OR "self-injury"[Title/Abstract] OR "self-harm"[Title/Abstract] OR "self-injury"[Title/Abstract] OR "psychological disorder"[Title/Abstract] OR "Psychiatric illness"[Title/Abstract] OR "psychiatric disorder*"[Title/Abstract]) OR ("Anxiety Disorders"[MeSH Terms] OR "Bipolar Disorder"[MeSH Terms] OR "Feeding and Eating Disorders"[MeSH Terms] OR "Depressive Disorder"[MeSH Terms] OR "Neurotic Disorders"[MeSH Terms] OR "Personality Disorders"[MeSH Terms] OR "Psychotic Disorders"[MeSH Terms] OR "Schizophrenia"[MeSH Terms] OR "Mental Disorders"[MeSH Terms:noexp] OR "Mental Health"[MeSH Terms] OR "Mentally Ill Persons"[MeSH Terms] OR "self-injurious behavior"[MeSH Terms] OR "psychology, clinical"[MeSH Terms]) OR ("Mental Health Services"[MeSH Terms]) OR ("mental health service*"[Title/Abstract] OR "CAMHS"[Title/Abstract] OR "psychiatry"[Title/Abstract] OR "psychology"[Title/Abstract] OR "psychotherap*"[Title/Abstract])) AND (("Telemedicine"[MeSH Terms:noexp] OR "Remote Consultation"[MeSH Terms] OR "Distance Counseling"[MeSH Terms] OR "therapy, computer-assisted"[MeSH Terms] OR "Videoconferencing"[MeSH Terms] OR "internet-based intervention"[MeSH Terms]) OR ("ehealth*"[Title/Abstract] OR "e health*"[Title/Abstract] OR "emedicine*"[Title/Abstract] OR "e medicine*"[Title/Abstract] OR "etherap*"[Title/Abstract] OR "e therap*"[Title/Abstract] OR "eCBT"[Title/Abstract] OR "e-CBT"[Title/Abstract] OR "iCBT"[Title/Abstract] OR "i-CBT"[Title/Abstract] OR "interap*"[all fields] OR "telemedicine"[Title/Abstract] OR "telecare"[Title/Abstract] OR "telepsychiatry"[Title/Abstract] OR "telecommunication*"[Title/Abstract] OR "teleconferencing"[Title/Abstract] OR "computer assisted therap*"[Title/Abstract] OR "teletherap*"[Title/Abstract] OR "telemental"[Title/Abstract] OR "e-mental health"[Title/Abstract] OR "e mental health"[Title/Abstract] OR "videoconferen*"[Title/Abstract] OR "video conferen*"[Title/Abstract]** OR “video-call*”[Title/Abstract] OR “Video call*”[Title/Abstract] or video-based call*[Title/Abstract] OR “video based call*”[Title/Abstract] **OR "videophone*"[Title/Abstract] OR "video phone*"[Title/Abstract]) OR (("mobile*"[Title/Abstract] OR "phone*"[Title/Abstract] OR "telephone*"[Title/Abstract] OR "remote*"[Title/Abstract] OR "distan*"[Title/Abstract] OR "online"[Title/Abstract] OR "virtual"[Title/Abstract] OR "electronic"[Title/Abstract] OR "email"[Title/Abstract] OR "e-mail"[Title/Abstract]) N2 ("consult*"[Title/Abstract] OR "counsel*"[Title/Abstract] OR "support*"[Title/Abstract] OR "interview*"[Title/Abstract] OR "monitor*"[Title/Abstract] OR "therap*"[Title/Abstract] OR "treatment*"[Title/Abstract] OR "CBT"[Title/Abstract])))**

*Limit to 2010 onwards.*

**Cochrane Systematic review database**

| 1 | MeSH descriptor: [Feeding and Eating Disorders] this term only |
| --- | --- |
| 2 | MeSH descriptor: [Anorexia Nervosa] this term only |
| 3 | MeSH descriptor: [Bulimia Nervosa] this term only |
| 4 | MeSH descriptor: [Binge-Eating Disorder] this term only |
| 5 | MeSH descriptor: [Bulimia] this term only |
| 6 | ("eating disorder*" or ` (eat* near/3 mood*) or EDNOS or anorexi* or orthorexi* or bulimi* or diabulimi* or (bing* near/2 (eat* or purg*))):ti,ab,kw |
| 7 | MeSH descriptor: [Mood Disorders] this term only |
| 8 | MeSH descriptor: [Depressive Disorder] this term only |
| 9 | MeSH descriptor: [Depressive Disorder, Major] this term only |
| 10 | MeSH descriptor: [Seasonal Affective Disorder] this term only |
| 11 | MeSH descriptor: [Dysthymic Disorder] this term only |
| 12 | MeSH descriptor: [Depression] this term only |
| 13 | (mood* or depress* or dysthymi* or "affective disorder*" or "affective symptom*"):ti,ab,kw |
| 14 | MeSH descriptor: [Anxiety Disorders] explode all trees |
| 15 | (general* near/2 anxi*):ti,ab,kw |
| 16 | anxiety:ti |
| 17 | ("anxiety disorder*" or "social* anxiety" or phobi* or agoraphobi* or anxious or obsess* or compulsi* or panic or PTSD or "post traumatic stress" or "posttraumatic stress" or neurosis or neuroses or neurotic):ti,ab,kw |
| 18 | ((psychological or emotional) near/2 (debrief* or stress* or trauma*)):ti,kw |
| 19 | MeSH descriptor: [Obsessive Behavior] this term only |
| 20 | MeSH descriptor: [Self-Injurious Behavior] explode all trees |
| 21 | ((self next (injur* or mutilat*)) or suicide* or suicidal or parasuicid* or para-suicid*):ti,ab,kw |
| 22 | MeSH descriptor: [Somatoform Disorders] explode all trees |
| 23 | ((conduct or behavi* or antisocial or anti-social or dyssocial or emotional* or internalizing or internalising or externalizing or externalising) near/2 (disorder* or problem* or difficult* or disturb* or psychopath*)):ti,ab,kw |
| 24 | MeSH descriptor: [Personality Disorders] explode all trees |
| 25 | (BPD or personality disorder*):ti,ab,kw |
| 26 | #1 OR #2 OR #3 OR #4 OR #5 OR #6 OR #7 OR #8 OR #9 OR #10 OR #11 OR #12 OR #13 OR #14 or #15 or #16 OR #17 OR #18 OR #19 OR #20 OR #21 OR #22 OR #23 OR #24 OR #25 |
| 27 | MeSH descriptor: [Dementia] explode all trees |
| 28 | MeSH descriptor: [Alzheimer Disease] 2 tree(s) exploded |
| 29 | MeSH descriptor: [Mental Health Services] explode all trees |
| 30 | MeSH descriptor: [Psychiatry] explode all trees |
| 31 | MeSH descriptor: [Psychotherapy] explode all trees |
| 32 | ("mental health service*" or CAMHS or "child and adolescent mental health service*" or psychiatry or psychology or psychotherap*):ti,ab,kw |
| 33 | #27 OR #28 OR #29 OR #30 OR #31 OR #32 |
| 34 | MeSH descriptor: [Telecommunications] explode all trees |
| 35 | MeSH descriptor: [Technology] this term only |
| 36 | MeSH descriptor: [Answering Services] explode all trees |
| 37 | MeSH descriptor: [Distance Counseling] explode all trees |
| 38 | MeSH descriptor: [Internet-Based Intervention] explode all trees |
| 39 | ((mobile* or phone* or telephone* or remote* or distan* or online or virtual or electronic or email or e-mail or video) N3 (consult* or counsel* or follow up or follow-up or support* or interview* or monitor* or therap* or treatment? or "CBT")):ti,ab,kw |
| 40 | (ehealth* or e-health* or emedicine* or e-medicine* or etherap* or e-therap* or eCBT or e-CBT or iCBT or i-CBT or Interap* or mhealth or m health or m-health or (electronic N2 CBT) or telemedicine or telecare or telepsychiatry or telecommunications or teleconferencing or computer assisted therap* or teletherap* or telemental or tele mental or tele-mental):ti,ab,kw |
| 41 | (videoconferen* or video-conferen* or videophone* Or video-phone* or video call* or video-call* or video call* or (video N2 call*) or (video-based N2 call*)):ti,ab,kw |
| 42 | #34 OR #35 OR #36 OR #37 OR #38 OR #39 OR #40 OR #41 |
| 43 | (#26 OR #33) AND #42 [limit to cochrane review database, 2010-2020] |

### Appendix 2a: Study Overlap

| Primary Study^a^ | Berryhill 2019a | Berryhill 2019b | Bolton 2015 | Christensen 2019 | Coughtrey 2018 | Dorstyn 2013 | Drago 2016 | Garcia-Lizana 2010 | Harerimana 2019 | Hassan 2019 | Lin 2019 | Lins 2014 | Muskens 2014 | Naslund 2020 | Norwood 2018 | Olthius 2016a | Othius 2016b | Turgoose 2018 | N reviews included |
| --- | --- | --- | --- | --- | --- | --- | --- | --- | --- | --- | --- | --- | --- | --- | --- | --- | --- | --- | --- |
| Aburizik 2013 |  |  |  |  |  |  |  |  | X |  |  |  |  |  |  |  |  |  | 1 |
| Acierno 2016 |  |  |  |  |  |  |  |  |  |  |  |  |  |  |  |  | X | X | 2 |
| Acierno 2017 |  |  |  |  |  |  |  |  |  |  |  |  |  |  |  |  |  | X | 1 |
| Ahmed 2008 |  |  |  |  |  |  |  |  |  |  |  |  |  | X |  |  |  |  | 1 |
| Amarendran 2011 |  |  |  |  |  |  | X |  |  |  |  |  |  |  |  |  |  |  | 1 |
| Andersson 2005 |  |  |  |  |  |  |  |  |  |  |  |  | X |  |  |  |  |  | 1 |
| Andersson 2009 |  |  |  |  |  |  |  |  |  |  |  |  |  |  |  | X |  |  | 1 |
| Andersson 2012 |  |  |  |  |  |  |  |  |  |  |  |  |  |  |  | X |  |  | 1 |
| Andersson 2012 |  |  |  |  |  |  |  |  |  |  |  |  |  |  |  | X |  |  | 1 |
| Andersson 2013 |  |  |  |  |  |  |  |  |  |  |  |  |  |  |  | X |  |  | 1 |
| Arnaert 2007 |  |  |  | X |  |  |  |  |  |  |  |  |  |  |  |  |  |  | 1 |
| Aziz 2004 |  |  |  |  |  |  |  |  |  |  |  |  | X |  |  |  |  |  | 1 |
| Baca 2007 |  |  |  |  |  |  |  |  |  |  | X |  |  |  |  |  |  |  | 1 |
| Barerra-Valencia 2017 |  |  |  |  |  |  |  |  |  |  |  |  |  | X |  |  |  |  | 1 |
| Berger 2009 |  |  |  |  |  |  |  |  |  |  |  |  |  |  |  | X |  |  | 1 |
| Berger 2011 |  |  |  |  |  |  |  |  |  |  |  |  |  |  |  | X |  |  | 1 |
| Berger 2014 |  |  |  |  |  |  |  |  |  |  |  |  |  |  |  | X |  |  | 1 |
| Bergstrom 2010 |  |  |  |  |  |  |  |  |  |  |  |  |  |  |  | X |  |  | 1 |
| Bishop 2002 |  |  |  | X |  |  |  | X |  | X |  |  |  |  |  |  |  |  | 3 |
| Bouchard 2004 |  | X |  |  |  |  |  | X |  | X |  |  |  |  | X |  |  |  | 4 |
| Brøndbo 2012 |  |  |  |  |  |  | X |  |  |  |  |  |  |  |  |  |  |  | 1 |
| Brooks 2013 |  |  |  |  |  |  |  |  |  |  |  |  |  |  |  |  |  | X | 1 |
| Burke 1995 |  |  |  |  |  |  |  |  |  |  |  |  | X |  |  |  |  |  | 1 |
| Butler 2012 |  |  |  |  |  |  |  |  |  |  |  |  |  | X |  |  |  |  | 1 |
| Cacciola 1999 |  |  |  |  |  |  |  |  |  |  |  |  | X |  |  |  |  |  | 1 |
| Carlbring 2004 |  |  |  |  |  |  |  |  |  |  |  |  |  |  |  | X |  |  | 1 |
| Carlbring 2006 |  |  |  |  |  |  |  |  |  |  |  |  |  |  |  | X |  |  | 1 |
| Carlbring 2007 |  |  |  |  |  |  |  |  |  |  |  |  |  |  |  | X |  |  | 1 |
| Carlson 2012 |  |  |  |  |  |  |  |  |  |  | X |  |  |  |  |  |  |  | 1 |
| Cernvall 2015 |  |  |  |  |  |  |  |  |  |  |  |  |  |  |  |  | X |  | 1 |
| Chang 1999 |  |  |  |  |  |  |  |  |  |  |  | X |  |  |  |  |  |  | 1 |
| Chang 2004 |  |  |  |  |  |  |  |  |  |  |  | X |  |  |  |  |  |  | 1 |
| Chang 2018 |  |  |  |  |  |  |  |  |  |  | X |  |  |  |  |  |  |  | 1 |
| Chiu 2009 |  |  |  |  |  | X |  |  |  |  |  |  |  |  |  |  |  |  | 1 |
| Choi 2012 |  |  |  |  |  | X |  |  |  |  |  |  |  |  |  |  |  |  | 1 |
| Choi 2014 |  |  |  |  |  |  |  | X |  |  |  |  |  |  |  |  |  |  | 1 |
| Choi 2014 |  |  |  |  |  |  | X |  | X |  |  |  |  |  |  |  |  |  | 2 |
| Choi 2014 |  |  |  | X |  |  | X |  |  |  |  |  |  |  |  |  |  |  | 2 |
| Chong 2012 |  |  |  |  |  | X |  |  |  | X |  |  |  |  |  |  |  |  | 2 |
| Clapp 2016 |  |  |  |  |  |  |  |  |  |  |  |  |  |  |  |  |  | X | 1 |
| Conn 2013 |  |  |  | X |  |  |  |  | X |  |  |  |  |  |  |  |  |  | 2 |
| Cowain 2001 |  | X |  |  |  |  |  |  |  |  |  |  |  |  |  |  |  |  | 1 |
| Crippa 2008 |  |  |  |  |  |  |  |  |  |  |  |  | X |  |  |  |  |  | 1 |
| Crowe 2016 |  |  |  | X |  |  |  |  |  |  |  |  |  |  |  |  |  |  | 1 |
| Davis 2004 |  |  |  |  |  |  |  |  |  |  |  | X |  |  |  |  |  |  | 1 |
| De Las Cuevas 2003 |  |  |  | X |  |  |  |  |  |  |  |  |  |  |  |  |  |  | 1 |
| De Las Cuevas 2006 |  |  |  |  |  |  | X | X |  | X |  |  |  |  |  |  |  |  | 3 |
| De Leo 2014 |  |  |  |  |  |  |  |  |  |  | X |  |  |  |  |  |  |  | 1 |
| Demiris 2011 |  | X |  |  |  |  |  |  |  |  |  |  |  |  |  |  |  |  | 1 |
| Dobkin 2011 |  |  |  |  | x |  |  |  |  |  |  |  |  |  |  |  |  |  | 1 |
| DuHamel 2010 |  |  |  |  |  |  |  |  |  |  |  |  |  |  |  |  | X |  | 1 |
| Dunstan 2012 |  | X |  |  |  |  |  |  |  |  |  |  |  |  |  |  |  |  | 1 |
| Dwight-Johnson 2011 |  |  |  |  | x | X |  |  |  |  |  |  |  |  |  |  |  |  | 2 |
| Egede 2016 |  |  |  | X |  |  |  |  |  |  |  |  |  |  |  |  |  |  | 1 |
| Egede 2017 |  |  |  |  |  |  |  |  |  |  |  |  |  | X |  |  |  |  | 1 |
| Eibl 2017 |  |  |  |  |  |  |  |  |  |  | X |  |  |  |  |  |  |  | 1 |
| Eisdorfer 2003 |  |  |  |  |  | X |  |  |  |  |  |  |  |  |  |  |  |  | 1 |
| Elford 2000 |  |  |  |  |  |  | X |  |  | X |  |  |  |  |  |  |  |  | 2 |
| Elford 2001 |  |  |  |  |  |  |  |  |  |  |  |  |  | X |  |  |  |  | 1 |
| Engel 2015 |  |  |  |  |  |  |  |  |  |  |  |  |  |  |  |  | X |  | 1 |
| Evans 2004 |  |  |  |  |  |  |  |  |  |  |  |  | X |  |  |  |  |  | 1 |
| Finkel 2007 |  |  |  |  |  |  |  |  |  |  |  | X |  |  |  |  |  |  | 1 |
| Fitt 2012 |  | X |  |  |  |  |  |  |  |  |  |  |  |  |  |  |  |  | 1 |
| Fortney 2007 |  |  |  | X |  |  | X |  |  |  |  |  |  |  |  |  |  |  | 2 |
| Fortney 2013 |  |  |  |  |  |  | X |  |  | X |  |  |  |  |  |  |  |  | 2 |
| Fortney 2015 |  |  |  |  |  |  |  |  |  |  |  |  |  |  |  |  |  | X | 1 |
| Frank 2017 |  |  |  | X |  |  |  |  |  |  |  |  |  |  |  |  |  |  | 1 |
| Franklin 2016 |  |  |  |  |  |  |  |  |  |  |  |  |  |  |  |  | X | X | 2 |
| Frueh 2007 |  |  |  |  |  |  |  | X |  | X |  |  |  |  |  |  | X | X | 4 |
| Frueh 2005 |  |  |  |  |  |  |  |  |  |  | X |  |  |  |  |  |  |  | 1 |
| Frueh 2007 |  |  |  |  |  |  |  | X |  |  |  |  |  |  |  |  |  |  | 1 |
| Furmark 2009 |  |  |  |  |  |  |  |  |  |  |  |  |  |  |  | X |  |  | 1 |
| Gant 2007 |  |  |  |  |  |  |  |  |  |  |  | X |  |  |  |  |  |  | 1 |
| Garzon-maldonado 2017 |  |  |  |  |  |  |  |  |  |  |  |  |  | X |  |  |  |  | 1 |
| Gerlach-Reinholz 2017 |  |  |  |  |  |  |  |  |  |  |  |  |  | X |  |  |  |  | 1 |
| Germain 2009 |  | X | X |  |  |  |  |  |  |  |  |  |  |  | X |  |  |  | 3 |
| Glueckauf 2012 |  |  |  |  |  | X |  |  |  |  |  | X |  |  |  |  |  |  | 2 |
| Godelski 2012 |  |  |  |  |  |  |  |  | X |  |  |  |  |  |  |  |  |  | 1 |
| Gonzalez 2015 |  |  |  |  |  |  |  |  |  |  |  |  |  |  |  |  |  |  | 0 |
| Greene 2010 |  |  |  |  |  |  |  |  |  |  |  |  |  |  |  |  |  | X | 1 |
| Greenwood 2004 |  |  |  | X |  |  |  |  |  |  |  |  |  |  |  |  |  |  | 1 |
| Griffiths 2006 |  | X |  |  |  |  |  |  |  |  |  |  |  |  |  |  |  |  | 1 |
| Gros 2011 |  | X | X |  |  |  |  |  |  |  |  |  |  |  |  |  |  | X | 3 |
| Gros 2012 |  |  |  |  |  |  |  |  |  |  |  |  |  |  |  |  |  |  | 0 |
| Gros 2016 |  |  |  |  |  |  |  |  |  |  |  |  |  |  |  |  |  | X | 1 |
| Grubbs 2015 |  |  |  |  |  |  |  |  |  |  |  |  |  |  |  |  |  | X | 1 |
| Grubbs 2017 |  |  |  |  |  |  |  |  |  |  |  |  |  |  |  |  |  | X | 1 |
| Hajebi 2012 |  |  |  |  |  |  |  |  |  |  |  |  | X |  |  |  |  |  | 1 |
| Hassija 2011 |  |  |  |  |  |  |  |  |  |  |  |  |  |  |  |  |  |  | 0 |
| Hedman 2011 |  |  |  |  |  |  |  |  |  |  |  |  |  |  |  | X |  |  | 1 |
| Hernandez-Tejada 2014 |  |  |  |  |  |  |  |  |  |  |  |  |  |  |  |  |  | X | 1 |
| Hilty 2007 |  |  |  | X |  |  |  |  |  | X |  |  |  |  |  |  |  |  | 2 |
| Himelhoch 2011 |  |  |  |  | x |  |  |  |  |  |  |  |  |  |  |  |  |  | 1 |
| Himle 2006 |  |  |  |  |  |  |  |  |  |  |  |  |  |  | X |  |  |  | 1 |
| Hull 2017 |  |  |  |  |  |  |  |  |  |  |  |  |  | X |  |  |  |  | 1 |
| Ivarsson 2014 |  |  |  |  |  |  |  |  |  |  |  |  |  |  |  | X | X |  | 2 |
| Jaconis 2017 |  |  |  |  |  |  |  |  |  |  |  |  |  |  |  |  |  | X | 1 |
| Jang 2014 |  |  |  | X |  | X |  |  |  |  |  |  |  |  |  |  |  |  | 2 |
| Johnston 2011 |  |  |  |  |  |  |  |  |  |  |  |  |  |  |  | X |  |  | 1 |
| Jones 2012 |  |  |  |  |  |  |  |  |  |  |  |  |  | X |  |  |  |  | 1 |
| Jones 2014 |  |  |  |  |  |  |  |  |  |  |  |  |  | X |  |  |  |  | 1 |
| Jong 2004 |  |  |  |  |  |  |  |  |  |  |  |  |  | X |  |  |  |  | 1 |
| Kennedy 2000 |  |  |  |  |  |  |  |  |  |  |  |  |  | X |  |  |  |  | 1 |
| Kim 2016 |  |  |  |  |  |  |  |  |  |  | X |  |  |  |  |  |  |  | 1 |
| King 2009 |  |  |  |  |  |  |  |  |  |  | X |  |  |  |  |  |  |  | 1 |
| King 2014 |  |  |  |  |  |  |  |  |  |  | X |  |  |  |  |  |  |  | 1 |
| Kiropoulos 2008 |  |  |  |  |  |  |  |  |  |  |  |  |  |  |  | X |  |  | 1 |
| Klee 2016 |  |  |  |  |  |  |  |  |  |  |  |  |  |  |  |  |  | X | 1 |
| Klein 2010 |  |  | X |  |  |  |  |  |  |  |  |  |  |  |  |  |  |  | 1 |
| Knaevelsrud 2015 |  |  |  |  |  |  |  |  |  |  |  |  |  |  |  |  | X |  | 1 |
| Kobak 2008 |  |  |  | X |  |  |  |  |  |  |  |  |  |  |  |  |  |  | 1 |
| Kobak 2015 |  |  |  |  |  |  |  |  |  |  |  |  |  |  |  | X |  |  | 1 |
| Kok 2014 |  |  |  |  |  |  |  |  |  |  |  |  |  |  |  | X |  |  | 1 |
| Lazzari 2011 |  |  |  |  |  |  |  |  |  |  |  |  |  |  |  |  |  |  | 0 |
| Lewis 2013 |  |  | X |  |  |  |  |  |  |  |  |  |  |  |  |  |  |  | 1 |
| Lichstein 2013 |  |  |  |  |  |  |  |  | X |  |  |  |  |  | X |  |  |  | 2 |
| Lightstone 2015 |  |  |  |  |  |  |  |  |  |  |  |  |  |  |  |  |  | X | 1 |
| Lindsay 2015 |  |  |  |  |  |  |  |  |  |  |  |  |  |  |  |  |  | X | 1 |
| Littleton 2012 |  |  | X |  |  |  |  |  |  |  |  |  |  |  |  |  |  |  | 1 |
| Litz 2007 |  |  | X |  |  |  |  |  |  |  |  |  |  |  |  |  | X |  | 2 |
| Lovell 2000 |  |  |  |  | X |  |  |  |  |  |  |  |  |  |  |  |  |  | 1 |
| Luxton 2015 |  | X |  |  |  |  |  |  |  |  |  |  |  |  |  |  |  | X | 2 |
| Luxton 2016 |  | X |  | X |  |  |  |  |  |  |  |  |  |  |  |  |  |  | 2 |
| Lyneham 2005 |  |  |  |  |  |  |  |  |  |  |  |  | X |  |  |  |  |  | 1 |
| Maieritsch 2015 |  |  |  |  |  |  | X |  |  |  |  |  |  |  |  |  | X | X | 3 |
| Malhotra 2014 |  |  |  |  |  |  | X |  |  |  |  |  |  |  |  |  |  |  | 1 |
| Manchanda 1998 |  |  |  |  |  |  |  |  |  |  |  |  |  |  | X |  |  |  | 1 |
| Manguno-Mire 2007 |  |  |  |  |  |  |  | X |  | X |  |  |  |  |  |  |  |  | 2 |
| Marchand 2011 |  | X | X |  |  |  |  |  |  |  |  |  |  |  |  |  |  |  | 2 |
| Matsuura 2000 |  |  |  |  |  |  | X |  |  |  |  |  |  |  |  |  |  |  | 1 |
| Mclellan 2017 |  | X |  |  |  |  |  |  |  |  |  |  |  |  |  |  |  |  | 1 |
| Menon 2001 |  |  |  | X |  |  |  |  |  |  |  |  |  |  |  |  |  |  | 1 |
| Miller 2002 |  |  |  |  | x |  |  |  |  |  |  |  |  |  |  |  |  |  | 1 |
| Miller 2016 |  |  |  |  |  |  |  |  |  |  |  |  |  |  |  |  |  | X | 1 |
| Mitchell 2008 |  |  |  |  |  |  | X | X |  | X |  |  |  |  | X |  |  |  | 4 |
| Modai 2006 |  |  |  |  |  |  |  |  |  |  |  |  |  | X |  |  |  |  | 1 |
| Mohr 2000 |  |  |  |  | x |  |  |  |  |  |  |  |  |  |  |  |  |  | 1 |
| Mohr 2005 |  |  |  |  | x |  |  |  |  |  |  |  |  |  |  |  |  |  | 1 |
| Mohr 2006 |  |  |  |  | x |  |  |  |  |  |  |  |  |  |  |  |  |  | 1 |
| Mohr 2011 |  |  |  |  | x |  |  |  |  |  |  |  |  |  |  |  |  |  | 1 |
| Mohr 2013 |  |  |  |  |  |  |  |  | X |  |  |  |  |  |  |  |  |  | 1 |
| Moreno 2012 |  |  |  |  |  | X | X |  |  |  |  |  |  |  |  |  |  |  | 2 |
| Morland 2004 |  |  |  |  |  |  |  |  |  |  |  |  |  |  |  |  |  | X | 1 |
| Morland 2010 |  |  |  |  |  |  | X |  |  |  |  |  |  |  |  |  |  | X | 2 |
| Morland 2011 |  |  |  |  |  |  |  |  |  |  |  |  |  |  |  |  |  | X | 1 |
| Morland 2013 |  |  | X |  |  |  |  |  |  |  |  |  |  | X |  |  |  |  | 2 |
| Morland 2014 |  |  |  |  |  |  | X |  |  |  |  |  |  |  |  |  | X | X | 3 |
| Morland 2015 |  |  |  |  |  |  |  |  |  |  |  |  |  |  |  |  |  | X | 1 |
| Morland 2015 |  |  |  |  |  |  |  |  |  |  |  |  |  |  |  |  |  | X | 1 |
| Morland 2015 |  |  |  |  |  |  |  |  |  |  |  |  |  |  | X |  | X | X | 3 |
| Munro Cullum 2014 |  |  |  |  |  |  | X |  |  |  |  |  |  |  |  |  |  |  | 1 |
| Nelson 2003 |  |  |  |  |  |  |  | X |  | X |  |  |  |  |  |  |  |  | 2 |
| Neufeld 2013 |  |  |  |  |  |  |  |  |  |  |  |  |  | X |  |  |  |  | 1 |
| Newby 2013 |  |  |  |  |  |  |  |  |  |  |  |  |  |  |  | X |  |  | 1 |
| Nieminen 2016 |  |  |  |  |  |  |  |  |  |  |  |  |  |  |  |  | X |  | 1 |
| Niles 2012 |  |  |  |  |  |  |  |  |  |  |  |  |  |  |  |  | X | X | 2 |
| Nordgren 2014 |  |  |  |  |  |  |  |  |  |  |  |  |  |  |  | X |  |  | 1 |
| O’Reilly 2007 |  |  |  | X |  |  |  | X |  | X |  |  |  |  |  |  |  |  | 3 |
| Ojserkis 2013 |  | X |  |  |  |  |  |  |  |  |  |  |  |  |  |  |  |  | 1 |
| Olthuis 2015 |  |  |  |  | X |  |  |  |  |  |  |  |  |  |  |  |  |  | 1 |
| Paing 2010 |  |  |  |  |  |  |  |  |  |  |  |  | X |  |  |  |  |  | 1 |
| Painter 2017 |  |  |  |  |  |  |  |  |  |  |  |  |  | X |  |  |  |  | 1 |
| Paulsen 1988 |  |  |  |  |  |  |  |  |  |  |  |  | X |  |  |  |  |  | 1 |
| Paxling 2011 |  |  |  |  |  |  |  |  |  |  |  |  |  |  |  | X |  |  | 1 |
| Poon 2005 |  |  |  |  |  |  | X |  |  |  |  |  |  |  |  |  |  |  | 1 |
| Price |  |  |  |  |  |  |  |  |  |  |  |  |  |  |  |  |  | X | 1 |
| Pyne 2010 |  |  |  |  |  |  |  |  |  |  |  |  |  | X |  |  |  |  | 1 |
| Rabinowitz 2010 |  |  |  |  |  |  |  |  |  |  |  |  |  | X |  |  |  |  | 1 |
| Ransom 2008 |  |  |  |  | x |  |  |  |  |  |  |  |  |  |  |  |  |  | 1 |
| Revicki 1997 |  |  |  |  |  |  |  |  |  |  |  |  | X |  |  |  |  |  | 1 |
| Richter 2015 |  |  |  |  |  |  |  |  |  |  | X |  |  |  |  |  |  |  | 1 |
| Robinson 2010 |  |  |  |  |  |  |  |  |  |  |  |  |  |  |  | X |  |  | 1 |
| Rohde 1997 |  |  |  |  |  |  |  |  |  |  |  |  | X |  |  |  |  |  | 1 |
| Ruskin 2004 |  |  |  |  |  |  | X | X |  | X |  |  |  | X |  |  |  |  | 4 |
| Russell 2015 |  |  |  |  |  |  |  |  | X |  |  |  |  |  |  |  |  |  | 1 |
| Salfi 2004 |  |  |  |  |  |  |  |  |  |  |  | X |  |  |  |  |  |  | 1 |
| Schutte 2015 |  |  |  |  |  |  | X |  |  |  |  |  |  |  |  |  |  |  | 1 |
| Seidel 2014 |  |  |  |  |  |  | X |  |  |  |  |  |  |  |  |  |  |  | 1 |
| Shealy 2015 |  | X |  |  |  |  |  |  |  |  |  |  |  |  |  |  |  |  | 1 |
| Shore 2007 |  |  |  |  |  |  | X |  |  |  |  |  |  |  |  |  |  |  | 1 |
| Shore 2012 |  |  |  |  |  |  |  |  |  |  |  |  |  |  |  |  |  | X | 1 |
| Shore 2014 |  |  |  | X |  |  |  |  |  |  |  |  |  |  |  |  |  |  | 1 |
| Simon 1993 |  |  |  |  |  |  |  |  |  |  |  |  | X |  |  |  |  |  | 1 |
| Simon 2009 |  |  |  |  |  |  |  |  |  |  |  |  |  | X |  |  |  |  | 1 |
| Simpson 2001 |  |  |  |  |  |  |  |  |  |  |  |  |  | X |  |  |  |  | 1 |
| Simpson 2001 |  |  |  | X |  |  |  |  |  |  |  |  |  |  |  |  |  |  | 1 |
| Simpson 2006 |  |  |  |  |  |  |  |  |  |  |  |  |  |  |  |  |  |  | 0 |
| Singh 2007 |  |  |  |  |  |  | X |  |  |  |  |  |  |  |  |  |  |  | 1 |
| Smith 2007 |  |  |  |  |  |  |  |  |  |  |  |  |  | X |  |  |  |  | 1 |
| Smolenski 2017 |  |  |  |  |  |  |  |  |  |  |  |  |  |  |  |  |  |  | 0 |
| Spaniel 2015 |  |  |  |  |  |  |  |  |  | X |  |  |  |  |  |  |  |  | 1 |
| Spaulding 2010 |  |  |  |  |  |  |  |  |  |  |  |  |  | X |  |  |  |  | 1 |
| Spence 2011 |  |  | X |  |  |  |  |  |  |  |  |  |  |  |  |  | X |  | 2 |
| Spence 2013 |  |  | X |  |  |  |  |  |  |  |  |  |  |  |  |  |  |  | 1 |
| Spence 2014 |  |  |  |  |  |  |  |  |  |  |  |  |  |  |  |  | X |  | 1 |
| Staton-Tindall 2014 |  |  |  |  |  |  |  |  |  |  | X |  |  |  |  |  |  |  | 1 |
| Stefan 2013 |  |  |  |  |  |  |  |  |  |  |  |  |  |  | X |  |  |  | 1 |
| Steffen 2000 |  |  |  |  |  |  |  |  |  |  |  | X |  |  |  |  |  |  | 1 |
| Stevens 1999 |  |  |  |  |  |  |  | X |  |  |  |  |  |  |  |  |  |  | 1 |
| Strachan 2012 |  | X |  |  |  |  |  |  |  |  |  |  |  |  |  |  | X | X | 3 |
| Stubbings 2013 |  | X |  |  |  |  |  |  |  |  |  |  |  |  | X |  |  |  | 2 |
| Swinson 1995 |  |  |  |  | X |  |  |  |  |  |  |  |  |  |  |  |  |  | 1 |
| Tan 2013 |  |  |  |  |  |  |  |  |  |  |  |  |  |  |  |  |  | X | 1 |
| Tang 2001 |  |  |  | X |  |  |  |  | X |  |  |  |  | X |  |  |  |  | 3 |
| Tarp 2017 |  |  |  |  |  |  |  |  |  |  | X |  |  |  |  |  |  |  | 1 |
| Taylor 2003 |  |  |  |  | X |  |  |  |  |  |  |  |  |  |  |  |  |  | 1 |
| Théberge-Lapointe 2015 |  | X |  |  |  |  |  |  |  |  |  |  |  |  |  |  |  |  | 1 |
| Thorp 2012 |  |  |  |  |  |  |  |  |  |  |  |  |  |  |  |  |  | X | 1 |
| Titov 2008 |  |  |  |  |  |  |  |  |  |  |  |  |  |  |  | X |  |  | 1 |
| Titov 2008 |  |  |  |  |  |  |  |  |  |  |  |  |  |  |  | X |  |  | 1 |
| Titov 2008 |  |  |  |  |  |  |  |  |  |  |  |  |  |  |  | X |  |  | 1 |
| Titov 2009 |  |  |  |  |  |  |  |  |  |  |  |  |  |  |  | X |  |  | 1 |
| Titov 2010 |  |  |  |  |  |  |  |  |  |  |  |  |  |  |  | X |  |  | 1 |
| Titov 2011 |  |  |  |  |  |  |  |  |  |  |  |  |  |  |  | X |  |  | 1 |
| Tremont 2008 |  |  |  |  |  |  |  |  |  |  |  | X |  |  |  |  |  |  | 1 |
| Tse 2015 |  |  |  |  |  |  | X |  |  |  |  |  |  |  |  |  |  |  | 1 |
| Tuerk 2010 |  |  | X |  |  |  |  |  |  |  |  |  |  |  |  |  |  | X | 2 |
| Tunstall 1997 |  |  |  |  |  |  |  |  |  |  |  |  | X |  |  |  |  |  | 1 |
| Tutty 2010 |  |  |  |  | x |  |  |  |  |  |  |  |  |  |  |  |  |  | 1 |
| Uebelacker 2011 |  |  |  |  |  | X |  |  |  |  |  |  |  |  |  |  |  |  | 1 |
| Vahia 2015 |  |  |  |  |  |  | X |  |  |  |  |  |  |  |  |  |  |  | 1 |
| Van Ballegooijen 2013 |  |  |  |  |  |  |  |  |  |  |  |  |  |  |  | X |  |  | 1 |
| van Bastelaar 2011 |  |  |  |  |  |  |  |  | X |  |  |  |  |  |  |  |  |  | 1 |
| Ward-King 2010 |  |  |  |  |  |  |  |  |  |  |  |  | X |  |  |  |  |  | 1 |
| Watson 1992 |  |  |  |  |  |  |  |  |  |  |  |  | X |  |  |  |  |  | 1 |
| Wells 1988 |  |  |  |  |  |  |  |  |  |  |  |  | X |  |  |  |  |  | 1 |
| Whealin 2015 |  |  |  |  |  |  |  |  |  |  |  |  |  |  |  |  |  | X | 1 |
| Wierwille 2016 |  |  |  |  |  |  |  |  |  |  |  |  |  |  |  |  |  | X | 1 |
| Wilz 2011 |  |  |  |  |  |  |  |  |  |  |  | X |  |  |  |  |  |  | 1 |
| Wims 2010 |  |  |  |  |  |  |  |  |  |  |  |  |  |  |  | X |  |  | 1 |
| Winter 2007 |  |  |  |  |  |  |  |  |  |  |  | X |  |  |  |  |  |  | 1 |
| Wray 2010 |  |  |  |  |  |  |  |  |  |  |  |  |  | X |  |  |  |  | 1 |
| Yeung 2009 |  |  |  | X |  |  |  |  |  |  |  |  |  |  |  |  |  |  | 1 |
| Yoshino 2001 |  |  |  |  |  |  | X |  |  |  |  |  |  |  |  |  |  |  | 1 |
| Yuen 2013 |  | X |  |  |  |  |  |  |  |  |  |  |  |  | X |  |  |  | 2 |
| Yuen 2015 |  | X |  |  |  |  |  |  |  |  |  |  |  |  |  |  | X | X | 3 |
| Zheng 2014 |  |  |  |  |  |  |  |  |  |  |  |  |  |  |  |  |  |  | 0 |
| Zheng 2017 |  |  |  |  |  |  |  |  |  |  | X |  |  |  |  |  |  |  | 1 |
| Ziemba 2014 |  | X | X |  |  |  |  |  |  |  |  |  |  |  |  |  | X | X | 4 |
| Note. Sansom Daley 2016's included papers are not listed here. The review included guidance only and so there was no study overlap.  ^a^ Refer to appendix 2b for full study reference | | | | | | | | | | | | | | | | | | |  |

### Appendix 2b: Primary studies full title

Aburizik 2013. A pilot randomized controlled trial of a depression and disease management programme delivered by phone.

Acierno 2016. Behavioural activation and therapeutic exposure for posttraumatic stress disorder: A noninferiority trial of treatment delivered in person versus home-based telehealth.

Acierno 2017. A non-inferiority trial of Prolonged Exposure for posttraumatic stress disorder: In person versus home-based telehealth.

Ahmed 2008. Feasibility of epilepsy follow-up care through telemedicine: a pilot study on the patient's perspective.

Amarendran 2011. The reliability of telepsychiatry for a neuropsychiatric assessment.

Andersson 2005. Internet-based self-help for depression: Randomised controlled trial

Andersson 2009. Internet-based self-help versus one-session exposure in the treatment of spider phobia: A randomized controlled trial.

Andersson 2012. Internet-based psychodynamic versus cognitive behavioural guided self-help for generalized anxiety disorder: A randomized controlled trial.

Andersson 2012. Therapist experience and knowledge acquisition in internet-delivered CBT for social anxiety disorder: a randomized controlled trial.

Andersson 2013. Internet-based exposure treatment versus one-session exposure treatment of snake phobia: A randomized controlled trial.

Arnaert 2007. Attitudes towards videotelephones: An exploratory study of older adults with depression.

Aziz 2004. Comparability of telephone and face-to-face interviews in assessing patients with posttraumatic stress disorder.

Baca 2007. Satisfaction with long-distance motivational interviewing for problem drinking.

Barerra-Valencia 2017. Cost-effectiveness of synchronous vs. asynchronous telepsychiatry in prison inmates with depression.

Berger 2009. Internet-based treatment for social phobia: A randomized controlled trial.

Berger 2011. Internet-based treatment of social phobia: A randomized controlled trial comparing unguided with two types of guided self-help.

Berger 2014. Internet-based guided self-help for several anxiety disorders: A randomized controlled trial comparing a tailored with a standardized disorder-specific approach.

Bergstrom 2010. Internet-versus group-administered cognitive behaviour therapy for panic disorder in a psychiatric setting: A randomised trial.

Bishop 2002. Client satisfaction in a feasibility study comparing face-to-face interviews with telepsychiatry.

Bouchard 2004. Delivering cognitive-behaviour therapy for panic disorder with agoraphobia in videoconference.

Brøndbo 2012. Agreement on web-based diagnoses and severity of mental health problems in Norwegian child and adolescent mental health services.

Brooks 2013. Reaching rural communities with culturally appropriate care: A model for adapting remote monitoring to American Indian veterans with posttraumatic stress disorder.

Burke 1995. The reliability and validity of the Geriatric Depression Rating Scale administered by telephone.

Butler 2012. Cost analysis of store-and-forward telepsychiatry as a consultation model for primary care.

Cacciola 1999. Comparability of telephone and in-person structured clinical interview for DSM-III-R (SCID) diagnoses.

Carlbring 2004. Treatment of panic disorder: Live therapy vs. self-help via the internet.

Carlbring 2006. Remote treatment of panic disorder: A randomized trial of internet-based cognitive behaviour therapy supplemented with telephone calls.

Carlbring 2007. Treatment of social phobia: Randomised trial of internet-delivered cognitive-behavioural therapy with telephone support.

Carlson 2012. Telehealth-delivered group smoking cessation for rural and urban participants: Feasibility and cessation rates.

Cernvall 2015. Internet-based guided self-help for parents of children on cancer treatment: A randomized controlled trial.

Chang 1999. Cognitive-behavioural intervention for homebound caregivers of persons with dementia.

Chang 2004. Perceived helpfulness of telephone calls.

Chang 2018. Expanding access to buprenorphine treatment in rural areas with telemedicine.

Chiu 2009. Internet-based care-giver support for Chinese Canadians taking care of a family member with Alzheimer disease and related dementia.

Choi 2012. culturally attuned Internet treatment for depression amongst Chinese Australians: a randomised controlled trial.

Choi 2014. Acceptance of home-based telehealth problem-solving therapy for depressed low income homebound older adults: qualitative interviews with the participants and aging-service case managers.

Choi 2014. Six month postintervention depression and disability outcomes of in-home telehealth problem-solving therapy for depressed, low-income homebound older adult.

Choi 2014. Telehealth problem-solving therapy for depressed low-income homebound older adults.

Chong 2012. Feasibility and acceptability of clinic-based telepsychiatry for low-income Hispanic primary care patients.

Clapp 2016. Patterns of change in response to prolonged exposure: Implications for treatment outcome.

Conn 2013. Program evaluation of a telepsychiatry service for older adults connecting a university-affiliated geriatric centre to a rural psychogeriatric outreach service in Northwest Ontario.

Cowain 2001. Cognitive-behavioural therapy via videoconferencing to a rural area.

Crippa 2008. Comparability between telephone and face-to-face structured clinical interview for DSM-IV in assessing social anxiety disorder.

Crowe 2016. A pilot program in rural telepsychiatry for deaf and hard of hearing populations.

Davis 2004. A comparison of in-home and telephone-based skill training interventions with caregivers of persons with dementia.

De Las Cuevas 2003. Telepsychiatry in the Canary Islands: User acceptance and satisfaction.

De Las Cuevas 2006. Randomized clinical trial of telepsychiatry through videoconference versus face-to-face conventional psychiatric treatment.

De Leo 2014. A brief behavioural telehealth intervention for veterans with alcohol misuse problems in VA primary care.

Demiris 2011. Use of videophones to deliver a cognitive-behavioural therapy to hospice caregivers.

Dobkin 2011. Telephone-based cognitive-behavioural therapy for depression in Parkinson disease.

DuHamel 2010. Randomized clinical trial of telephone-administered cognitive-behavioural therapy to reduce post-traumatic stress disorder and distress symptoms after hematopoietic stem-cell transplantation.

Dunstan 2012. Treatment via videoconferencing: a pilot study of delivery by clinical psychology trainees.

Dwight-Johnson 2011. Telephone-based cognitive-behavioural therapy for Latino patients living in rural areas: a randomized pilot study.

Egede 2016. Psychotherapy for depression in older veterans via telemedicine: Effect on quality of life, satisfaction, treatment credibility, and service delivery perception.

Egede 2017. Trajectory of cost overtime after psychotherapy for depression in older veterans via telemedicine.

Eibl 2017. The effectiveness of telemedicine-delivered opioid agonist therapy in a supervised clinical setting.

Eisdorfer 2003. The effect of a family therapy and technology-based intervention on caregiving depression.

Elford 2000. A randomized, controlled trial of child psychiatric assessments conducted using videoconferencing.

Elford 2001. A prospective satisfaction study and cost analysis of a pilot child telepsychiatry service in Newfoundland.

Engel 2015. Delivery of self-training and education for stressful situations(DESTRESS-PC): A randomized trial of nurse assisted online self-management for PTSD in primary care.

Evans 2004. Assessing mental health in primary care research using standardized scales: can it be carried out over the telephone?

Finkel 2007. E-care: a telecommunications technology intervention for family caregivers of dementia patients.

Fitt 2012. Metacognitive therapy for obsessive compulsive disorder by videoconference: a preliminary study.

Fortney 2007. A randomized trial of telemedicine-based collaborative care for depression.

Fortney 2013. Practice-based versus telemedicine-based collaborative care for depression in rural federally qualified health centres: a pragmatic randomized comparative effectiveness trial.

Fortney 2015. Telemedicine based collaborative care for posttraumatic stress disorder: a randomized clinical trial

Frank 2017. Video conference-based psychotherapeutic follow-up treatment. qualitative case study using CBASP approach.

Franklin 2016. Face to face but not in the same place: A pilot study of prolonged exposure therapy.

Frueh 2007. A randomized trial of telepsychiatry for post-traumatic stress disorder.

Frueh 2005. Telehealth service delivery for persons with alcoholism.

Frueh 2007. Therapist adherence and competence with manualized cognitive-behavioural therapy for PTSD delivered via videoconferencing technology.

Furmark 2009. Guided and unguided self-help for social anxiety disorder: Randomised controlled trial.

Gant 2007. Comparative outcomes of two distance-based interventions for male caregivers of family members with dementia.

Garzon-maldonado 2017. An assessment of telephone assistance systems for caregivers of patients with Alzheimer’s disease.

Gerlach-Reinholz 2017. Telefoncoaching bei depression [telephone coaching for depression].

Germain 2009. Effectiveness of cognitive behavioural therapy administered by videoconference for post-traumatic stress disorder.

Glueckauf 2012. Telephone-based, cognitive-behavioural therapy for African American dementia caregivers with depression: initial findings.

Godelski 2012. Home telemental health implementation and outcomes using electronic messaging.

Gonzalez 2015. Telehealth videoconferencing psychotherapy in rural primary care.

Greene 2010. How does tele-mental health affect group therapy process? Secondary analysis of a noninferiority trial.

Greenwood 2004. Evaluation of a rural telepsychiatry service.

Griffiths 2006. Telemedicine as a means of delivering cognitive-behavioural therapy to rural and remote mental health clients.

Gros 2011. Exposure therapy for PTSD delivered to veterans via telehealth: predictors of treatment completion and outcome and comparison to treatment delivered in person.

Gros 2012. Behavioural activation and therapeutic exposure: An investigation of relative symptom changes in PTSD and depression during the course of integrated behavioural activation, situational exposure, and imaginal exposure techniques.

Gros 2016. Treatment satisfaction of home-based telehealth versus in person delivery of prolonged exposure for combat-related PTSD in veterans.

Grubbs 2015. Predictors of initiation and engagement of cognitive processing therapy among veterans with PTSD enrolled in collaborative care.

Grubbs 2017. Usual care for rural veterans with posttraumatic stress disorder.

Hajebi 2012. Telephone versus face-to-face administration of the structured clinical interview for diagnostic and statistical manual of mental disorders, fourth edition, for diagnosis of psychotic disorders.

Hassija 2011. The effectiveness and feasibility of videoconferencing technology to provide evidence-based treatment to rural domestic violence and sexual assault populations.

Hedman 2011. Internet-based cognitive behaviour therapy vs. cognitive behavioural group therapy for social anxiety disorder: A randomized controlled non-inferiority trial.

Hernandez-Tejada 2014. Early treatment withdrawal from evidence-based psychotherapy for PTSD: Telemedicine and in-person parameters.

Hilty 2007. A randomized controlled trial of disease management modules, including telepsychiatric care for depression in rural primary care.

Himelhoch 2011. Feasibility of telephone- based cognitive behavioural therapy targeting major depression among urban dwelling African-American people with co-occurring HIV.

Himle 2006. Videoconferencing-based cognitive-behavioural therapy for obsessive-compulsive disorder.

Hull 2017. A study of asynchronous mobile-enabled SMS text psychotherapy.

Ivarsson 2014. Guided internet-delivered cognitive behaviour therapy for post-traumatic stress disorder: A randomized controlled trial.

Jaconis 2017. Concurrent treatment of PTSD and alcohol use disorder via telehealth in a female Iraq veteran.

Jang 2014. Telecounselling for the linguistically isolated: a pilot study with older Korean immigrants

Johnston 2011. A RCT of a transdiagnostic internet-delivered treatment for three anxiety disorders: Examination of support roles and disorder specific outcomes.

Jones 2012. Acceptability and cost-effectiveness of military telehealth mental health screening.

Jones 2014. Technology-enhanced program for child disruptive behaviour disorders: development and pilot randomized control trial.

Jong 2004. Managing suicides via videoconferencing in a remote northern community in Canada.

Kennedy 2000. A community-based approach to evaluation of health outcomes and costs for telepsychiatry in a rural population.

Kim 2016. A randomized controlled trial of a videoconferencing smoking cessation intervention for Korean American women: Preliminary findings.

King 2009. Assessing the effectiveness of an Internet-based videoconferencing platform for delivering intensified substance abuse counselling.

King 2014. A randomized trial of web-based videoconferencing for substance abuse counselling.

Kiropoulos 2008. Is internet-based CBT for panic disorder and agoraphobia as eGective as face-to-face CBT?

Klee 2016. Interest in technology based therapies hampered by access: A survey of veterans with serious mental illnesses.

Klein 2010. A therapist-assisted cognitive behaviour therapy for posttraumatic stress disorder: pre-, post-and 3-month follow-up results from an open trial.

Knaevelsrud 2015. Web-based psychotherapy for posttraumatic stress disorder in war-traumatized Arab patients: Randomized controlled trial.

Kobak 2008. Face-to-face versus remote administration of the Montgomery-Asberg Depression Rating Scale using videoconference and telephone.

Kobak 2015. Computer-assisted cognitive behaviour therapy for obsessive compulsive disorder: A randomized trial on the impact of lay vs. professional coaching.

Kok 2014. Short term effectiveness of web-based guided self-help for phobic outpatients: Randomized controlled trial.

Lazzari 2011. Behavioural activation treatment for depression in older adults delivered via videoconferencing: A pilot study.

Lewis 2013. Development of a guided self-help (GSH) program for the treatment of mild to-moderate Posttraumatic Stress Disorder (PTSD).

Lichstein 2013. Telehealth cognitive behaviour therapy for co-occurring insomnia and depression symptoms in older adults in the united states.

Lightstone 2015. Collaborative music therapy via remote video technology to reduce a veteran’s symptoms of severe, chronic PTSD.

Lindsay 2015. Implementation of video telehealth to improve access to evidence-based psychotherapy for posttraumatic stress disorder.

Littleton 2012. From survivor to thriver: a pilot study of an online program for rape victims.

Litz 2007. A randomized, controlled proof of concept trial of an internet-based therapist assisted self-management treatment for posttraumatic stress disorder.

Lovell 2000. Telephone treatment of obsessive-compulsive disorder.

Luxton 2015. An evaluation of the feasibility and safety of a home-based telemental health treatment for post-traumatic stress in the U.S. Military.

Luxton 2016. Home-based tele behavioural health for US military personnel and veterans with depression: A RCT.

Lyneham 2005. Agreement between telephone and in-person delivery of a structured interview for anxiety disorders in children.

Maieritsch 2015. Randomized controlled equivalence trial comparing videoconference and in person delivery of cognitive processing therapy for PTSD.

Malhotra 2014. Development of a novel diagnostic system for a tele psychiatric application: a pilot validation study.

Manchanda 1998. Cognitive behaviour therapy via interactive video.

Manguno-Mire 2007. The use of telemedicine to evaluate competency to stand trial: a preliminary randomized controlled study.

Marchand 2011. Relative efficacy of cognitive-behavioural therapy administered by videoconference for posttraumatic stress disorder: A six-month follow-up.

Matsuura 2000. Application of telepsychiatry: a preliminary study.

Mclellan 2017. Delivery of a therapist-facilitated telecare anxiety program to children in rural communities: a pilot study.

Menon 2001. Evaluation of a portable low-cost videophone system in the assessment of depressive symptoms and cognitive function in elderly mentally ill veterans.

Miller 2002. Interpersonal psychotherapy delivered over the telephone to recurrent depressives: A pilot study.

Miller 2016. Interest in use of technology for healthcare among veterans receiving treatment for mental health.

Mitchell 2008. A randomized trial comparing the efficacy of cognitive-behavioural therapy for bulimia nervosa delivered via telemedicine versus face-to-face.

Modai 2006. Cost-effectiveness, safety, and satisfaction with video telepsychiatry versus face-to-face care in ambulatory settings.

Mohr 2000. Telephone administered cognitive-behavioural therapy for the treatment of depressive symptoms in multiple sclerosis.

Mohr 2005. Telephone-administered psychotherapy for depression.

Mohr 2006. Telephone administered cognitive behavioural therapy for the treatment of depression in a rural primary care clinic.

Mohr 2011. Telephone-administered cognitive behavioural therapy for veterans served by community-based outpatient clients.

Mohr 2013. A randomized controlled trial evaluating a manualized TeleCoaching protocol for improving compliance with a web-based intervention for the treatment of depression.

Moreno 2012. Use of standard webcam and Internet equipment for tele-psychiatry treatment of depression among underserved Hispanics.

Morland 2004. Telemedicine and coping skills groups for Pacific Island veterans with 584 Journal of Telemedicine and Telecare.

Morland 2010. Telemedicine for anger management therapy in a rural population of combat veterans with posttraumatic stress disorder: a randomized noninferiority trial.

Morland 2011. Group cognitive processing therapy delivered to veterans via telehealth: A pilot cohort.

Morland 2013. Telemedicine: a cost-reducing means of delivering psychotherapy to rural combat veterans with PTSD.

Morland 2014. Cognitive processing therapy for posttraumatic stress disorder delivered to rural veterans via telemental health: a randomized noninferiority clinical trial.

Morland 2015. Telemedicine versus in-person delivery of cognitive processing therapy for women with posttraumatic stress disorder: A randomized noninferiority trial.

Morland 2015. Telemedicine versus in‐person delivery of cognitive processing therapy for women with posttraumatic stress disorder: A randomized noninferiority trial.

Morland 2015. Telemedicine versus in-person delivery of cognitive processing therapy for women with posttraumatic stress disorder: A randomized noninferiority trial.

Munro Cullum 2014. Teleneuropsychology: evidence for video teleconference-based neuropsychological assessment.

Nelson 2003. Treating childhood depression over videoconferencing.

Neufeld 2013. Walk-in telemental health clinics improve access and efficiency: a 2-year follow-up analysis.

Newby 2013. Internet cognitive behavioural therapy for mixed anxiety and depression: A randomized controlled trial and evidence of effectiveness in primary care.

Nieminen 2016. Internet-provided cognitive behaviour therapy of posttraumatic stress symptoms following childbirth – A randomized controlled trial.

Niles 2012. Comparing mindfulness and psychoeducation treatments for combat-related PTSD using a telehealth approach.

Nordgren 2014. Effectiveness and cost-effectiveness of individually tailored Internet-delivered cognitive behaviour therapy for anxiety disorders in a primary care population: A randomized controlled trial.

O’Reilly 2007. Is telepsychiatry equivalent to face-to-face psychiatry? Results from a randomized controlled equivalence trial.

Ojserkis 2013. Paediatric obsessive-compulsive disorder.

Olthuis 2015. Telephone delivered cognitive behavioural therapy for high anxiety sensitivity: A randomised controlled trial.

Paing 2010. Face-to-face versus telephone administration of the parent’s version of the children’s interview for psychiatric syndromes (P-ChIPS).

Painter 2017. Cost-effectiveness of telemedicine-based collaborative care for post-traumatic stress disorder.

Paulsen 1988. Reliability of the telephone interview in diagnosing anxiety disorders.

Paxling 2011. Guided internet-delivered cognitive behaviour therapy for generalized anxiety disorder: A randomized controlled trial.

Poon 2005. Cognitive intervention for community dwelling older persons with memory problems: telemedicine versus face-toface treatment.

Price. Examination of prior experience with telehealth and comfort with telehealth technology as a moderator of treatment response for PTSD and depression in veterans.

Pyne 2010. Cost-effectiveness analysis of a rural telemedicine collaborative care intervention for depression.

Rabinowitz 2010. Benefits of a telepsychiatry consultation service for rural nursing home residents.

Ransom 2008. Telephone delivered, interpersonal psychotherapy for HIV-infected rural persons with depression: A pilot trial.

Revicki 1997. Telephone versus in-person clinical and health status assessment interviews in patients with bipolar disorder.

Richter 2015. Comparative and cost effectiveness of telemedicine versus telephone counselling for smoking cessation.

Robinson 2010. Internet treatment for generalized anxiety disorder: A randomized controlled trial comparing clinician vs. technician assistance.

Rohde 1997. Comparability of telephone and face-to-face interviews in assessing axis I and II disorders.

Ruskin 2004. Treatment outcomes in depression: comparison of remote treatment through telepsychiatry to in-person treatment.

Russell 2015. Exploring the predictors of home telehealth uptake by elderly Australian healthcare consumers.

Salfi 2004. Seeking to understand telephone support for dementia caregivers: a qualitative case study.

Schutte 2015. Usability and reliability of a remotely administered adult autism assessment, the autism diagnostic observation schedule (ADOS) module 4.

Seidel 2014. Agreement between telepsychiatry assessment and face-to-face assessment for Emergency Department psychiatry patients.

Shealy 2015. Delivering an evidence-based mental health treatment to underserved populations using telemedicine: the case of a trauma-affected adolescent in a rural setting.

Shore 2007. Diagnostic reliability of telepsychiatry in American Indian veterans.

Shore 2012. Characteristics of telemental health service use by American Indian veterans.

Shore 2014. Meeting veterans where they're @: a VA-based Telemental Health (HBTMH) pilot program.

Simon 1993. Telephone assessment of depression severity.

Simon 2009. Incremental benefit and cost of telephone care management and telephone psychotherapy doe depression in primary care.

Simpson 2001. Evaluation of a routine telepsychiatry service.

Simpson 2001. Telepsychiatry as a routine service-the perspective of the patient.

Simpson 2006. Does video therapy work? A single case series of bulimic disorders.

Singh 2007. Accuracy of telepsychiatric assessment of new routine outpatient referrals.

Smith 2007. A cost-minimization analysis of a telepaediatric mental health service for patients in rural and remote Queensland.

Smolenski 2017. Unobserved heterogeneity in response to treatment for depression through videoconference.

Spaniel 2015. Psychiatrist's adherence: a new factor in relapse prevention in schizophrenia A randomized controlled study on relapse control through telemedicine system.

Spaulding 2010. Cost savings of telemedicine utilization for child psychiatry in a rural Kansas community.

Spence 2011. Randomized controlled trial of internet-delivered cognitive behavioural therapy for Posttraumatic Stress Disorder.

Spence 2013. Internet-delivered eye movement desensitization and reprocessing: and open trial

Spence 2014. Internet-based trauma-focused cognitive behavioural therapy for PTSD with and without exposure components: A randomized controlled trial.

Staton-Tindall 2014. METelemedicine: A pilot study with rural alcohol users on community supervision.

Stefan 2013. Face‐to‐face counselling versus high definition holographic projection system. Efficacy and therapeutic alliance. A brief research report.

Steffen 2000. Anger management for dementia caregivers: a preliminary study using video and telephone interventions.

Stevens 1999. Pilot study of televideo psychiatric assessments in an underserviced community.

Strachan 2012. An integrated approach to delivering exposure-based treatment for symptoms of PTSD and depression in OIF/OEF veterans: preliminary findings.

Stubbings 2013. Comparing in-person to videoconference-based cognitive behavioural therapy for mood and anxiety disorders: randomized controlled trial.

Swinson 1995. Efficacy of telephone- administered behavioural therapy for panic disorder with agoraphobia.

Tan 2013. Improving access to care for women veterans suffering from chronic pain and depression associated with trauma.

Tang 2001. Telepsychiatry in psychogeriatric service: a pilot study.

Tarp 2017. Effectiveness of optional videoconferencing-based treatment of alcohol use disorders: Randomized controlled trial.

Taylor 2003. Telephone-administered cognitive-behaviour therapy for obsessive-compulsive disorder.

Théberge-Lapointe 2015. Efficacy of a cognitive-behavioural therapy administered by videoconference for generalized anxiety disorder.

Thorp 2012. Lessons learned from studies of psychotherapy for posttraumatic stress disorder via video teleconferencing.

Titov 2008. Shyness 1: Distance treatment of social phobia over the internet.

Titov 2008. Shyness 2: Treating social phobia online: Replication and extension.

Titov 2008. Shyness 3: Randomized controlled trial of guided versus unguided internet-based CBT for social phobia.

Titov 2009. Clinician-assisted internet based treatment is effective for generalized anxiety disorder: Randomized controlled trial.

Titov 2010. Transdiagnostic internet treatment for anxiety disorders: A randomized controlled trial.

Titov 2011. Transdiagnostic internet treatment for anxiety and depression: A randomised controlled trial.

Tremont 2008. Telephone delivered psychosocial intervention reduces burden in dementia caregivers.

Tse 2015. Teletherapy delivery of caregiver behaviour training for children with attention-deficit hyperactivity disorder.

Tuerk 2010. A pilot study of prolonged exposure therapy for Posttraumatic Stress Disorder delivered via telehealth technology.

Tunstall 1997. Concurrent validity of a telephone-administered version of the Gospel Oak instrument (including the SHORT-CARE).

Tutty 2010. Evaluating the effectiveness of cognitive-behavioural teletherapy in depressed adults.

Uebelacker 2011. Telephone depression care management for Latino Medicaid health plan members: a pilot randomized controlled trial.

Vahia 2015. Telepsychiatry for neurocognitive testing in older rural Latino adults.

Van Ballegooijen 2013. An Internet-based guided self-help intervention for panic symptoms: Randomized controlled trial.

van Bastelaar 2011. Web-based depression treatment for type 1 and type 2 diabetic patients: a randomized, controlled trial.

Ward-King 2010. Brief report: telephone administration of the autism diagnostic interview– revised: reliability and suitability for use in research.

Watson 1992. Comparability of telephone and face to face diagnostic interview schedules.

Wells 1988. Agreement between face-to-face and telephone-administered versions of the depression section of the NIMH diagnostic interview schedule.

Whealin 2015. E-mental health preferences of veterans with and without probable posttraumatic stress disorder.

Wierwille 2016. Effectiveness of PTSD telehealth treatment in a VA clinical sample.

Wilz 2011. Goal attainment and treatment compliance in a cognitive-behavioural telephone intervention for family caregivers of persons with dementia.

Wims 2010. Clinician-assisted internet-based treatment is effective for panic: A randomized controlled trial.

Winter 2007. Evaluation of a telephone-based support group intervention for female caregivers of community dwelling individuals with dementia.

Wray 2010. The effect of telephone support groups on costs of care for veterans with dementia.

Yeung 2009. Feasibility and effectiveness of telepsychiatry services for Chinese immigrants in a nursing home.

Yoshino 2001. Telepsychiatry: assessment of televideo psychiatric interview reliability with present- and next-generation Internet infrastructures.

Yuen 2013. Acceptance based behaviour therapy for social anxiety disorder through videoconferencing.

Yuen 2015. Randomized controlled trial of home-based telehealth versus in-person prolonged exposure for combat-related PTSD in veterans: preliminary results.

Zheng 2014. Telehealth-based therapy connecting rural Mandarin-speaking traumatized clients with a Mandarin-speaking therapist.

Zheng 2017. Treatment outcome comparison between telepsychiatry and face-to-face buprenorphine medication-assisted treatment for opioid use disorder: A 2-year retrospective data analysis.

Ziemba 2014. Posttraumatic stress disorder treatment for operation enduring freedom/operation Iraqi freedom com-bat veterans through a civilian community-based telemedicine network.

### Appendix 3: Quality assessment further detail

| AMSTAR2 Criteria | Harerimana 2019 | Dorstyn 2013 | Berryhill 2019a | Berryhill 2019b | Bolton 2015 | Christensen 2019 | Coughtrey 2018 | Drago 2016 | Garcia-Lizana 2010 | Hassan 2019 | Lin  2019 | Lins  2014 | Muskens 2014 | Norwood 2018 | Olthuis 2016a | Olthuis 2016b | Sansom-Daly 2016 | Turgoose 2018 |
| --- | --- | --- | --- | --- | --- | --- | --- | --- | --- | --- | --- | --- | --- | --- | --- | --- | --- | --- |
| 1 | Yes | Yes | Yes | Yes | Yes | yes | Yes | Yes | Yes | Yes | Yes | Yes | Yes | Yes | Yes | Yes | No | No |
| 2 | Yes | No | No | No | No | No | No | No | No | No | No | Partial yes | No | No | Yes | No | No | No |
| 3 | Yes | No | No | No | Yes | Yes | Yes | Yes | No | Yes | Yes | Yes | Yes | Yes | Yes | Yes | Yes | Yes |
| 4 | No | Partial yes | No | No | Yes | Partial yes | Yes | Yes | Partial yes | Yes | Yes | Yes | Yes | Yes | Partial yes | Partial yes | Yes | No |
| 5 | No | No | No | No | No | No | No | No | Yes | No | No | Yes | Yes | No | Yes | No | Yes | No |
| 6 | No | No | Yes | Yes | No | Yes | Yes | Yes | Yes | Yes | Yes | Partial yes | Yes | No | Yes | Yes | No | No |
| 7 | No | No | No | No | No | No | No | No | No | No | No | Yes | No | No | Yes | No | No | No |
| 8 | Yes | Partial yes | Yes | Yes | Yes | Partial yes | Partial yes | Yes | Partial yes | Partial yes | Yes | Partial yes | Partial yes | Partial yes | Yes | Partial yes | Yes | Yes |
| 9 | Yes | No | Yes | Yes | No | Partial yes | Yes | Partial yes | No | No | Yes | Yes | Yes | yes | Yes | Yes | Yes | No |
| 10 | No | No | No | No | No | No | Yes | Yes | No | No | No | Yes | No | No | No | No | No | No |
| 11 | No MA | No MA | No MA | No MA | Yes | No MA | No MA | Yes | No MA | No MA | No MA | Yes | No MA | Yes | Yes | Yes | No MA | No MA |
| 12 | No MA | No MA | No MA | No MA | Partial yes | No MA | No MA | No | No MA | No MA | No MA | Yes | No MA | Yes | No | No | No MA | No MA |
| 13 | No | No | No | No | No | No | No | No | No | No | Yes | Yes | Yes | yes | Yes | Yes | No | No |
| 14 | Yes | No | No | No | No | No | Yes | Yes | No | No | Yes | Yes | Yes | Yes | Yes | Yes | Yes | Yes |
| 15 | No MA | No MA | No MA | No MA | Yes | No MA | No MA | Yes | No MA | No MA | No MA | No | No MA | No | Yes | Yes | No MA | No MA |
| 16 | Yes | No | Yes | Yes | Yes | Yes | Yes | Yes | Yes | Yes | Yes | Yes | Yes | Yes | Yes | No | Yes | Yes |
| *MA: Meta-Analysis* | | | | | | | | | | | | | | | | | | |
| *1. PICO criteria included*  *2. Explicit statement that the review methods were established prior to the conduct of the review*  *3 Selection of study designs to include explained*  *4. Comprehensive literature search strategy*  *5.Study selection performed in duplicate*  *6. Data extraction performed in duplicate*  *7. List of excluded studies and reasons provided*  *8. Included studies described in adequate detail*  *9.Satisfactory assessment of risk of bias (RoB) in individual studies*  *10. Sources of funding reported*  *11. Meta-Analysis: appropriate methods for statistical combination of results*  *12. Meta-Analysis: Assessment of the potential impact of RoB in individual studies*  *13. Interpreation accounts for RoB*  *14. Satisfactory explanation for, and discussion of, any heterogeneity observed in the results of the review*  *15. Meta-Analysis: Adequate investigation of publication bias*  *16. Potential conflicts of interest reported* | | | | | | | | | | | | | | | | | | |

### Appendix 4: Guideline recommendations (Sansom-Daley 2016)

|  | Guidelines |
| --- | --- |
| *Appropriateness of e-mental health* | Client related factors: Firm recommendations that mental health professionals should incorporate an assessment process to determine the appropriateness of e-mental health services for an individual client (58%, 11/19 guidelines). However, only four sets of guidelines provided more concrete recommendations as to how professionals could undertake such an assessment Service related factors: Tentative recommendations that psychological tests designed to be implemented face to face may not be possible or ethical to conduct online. |
| *Competence* | Competence:  1) Firm recommendations that mental health professionals should provide online services within the boundaries of their competence, with an understanding of the limits and applications of different technologies. (58%, 11/19 guidelines).  2) Firm recommendations that professionals should acquire skills to manage technology they are using (53%, 10/19 guidelines).  3) Tentative recommendations that mental health professionals should be culturally competent to deliver online services to different populations, including considerations of clients' ethnic/racial, cultural, linguistic, gender/sexual orientation, geographic, and socioeconomic backgrounds. (42%, 8/19 guidelines). |
| *Legal and regulatory issues* | 1) Firm recommendations that professionals should know and comply with all relevant laws and regulations (79%, 15/19 guidelines) 2) Firm recommendations that professionals should ensure that their licensing board approves of the provision of online services, and obtain site-specific credentialing across jurisdictions where necessary (63%, 12/19)  3) Tentative recommendations that professionals delivering e-mental health interventions should adhere to the usual laws and professional standards applicable to record keeping, particularly where the intervention diverges from usual practice (47%, 9/19 guidelines).  4) Tentative recommendations that professionals should take steps to determine the age of potential clients to establish the appropriateness of e-mental health interventions, and should ensure that a parent/guardian’s consent is obtained for all minors before services proceed (21%, 4/19 guidelines) |
| *Confidentiality* | 1) Firm recommendation that mental health professionals should take all up to date precautionary efforts to protect clients' confidentiality using e-mental health services (63%, 12/19 guidelines).  2) Tentative recommendations that privacy during sessions, anonymity, and identity should be ensured in the use and storage of electronic materials |
| *Consent* | 1) Firm recommendations that documenting thorough consent processes consistent with relevant laws and regulations is important (58%, 11/19 guidelines) 2) Firm recommendations that these consent processes should address numerous issues unique to e-mental health services including privacy and confidentiality in the online domain, security steps taken, technological equipment and skills requirements, limits to communication, and reliability of the connection (63%, 12/19 guidelines)  3) Tentative recommendations that professionals should clarify contact information, and the nature of and expectations around therapeutic contact at the commencement of e-mental health interventions (47%, 9/19 guidelines) 4) Tentative recommendations that professionals should clarify expected timeframes for their client receiving a response from them, as well as processes around emergency contacts (47%, 9/19 guidelines) |
| *Professional boundaries* | 1) Tentative recommendations that mental health professionals should consider the increased potential for boundary issues to arise using e-mental health (21%, 4/19 guidelines)  2) Tentative recommendations that professionals should use the same level of professional language across all media as they would in person (21%, 4/19 guidelines) |
| *Crisis intervention and distress management* | 1) Firm recommendations that mental health professionals should establish in-person clinical supports in the client's geographic location prior to initiating e-mental health services, in case of emergency (53%, 10/19 guidelines) 2) Tentative recommendations that professionals should inform clients of alternative means of communication should the technology fail (42%, 8/19 guidelines) 3) Tentative recommendations that professionals should be familiar with mandatory reporting and involuntary hospitalisation laws (21%, 4/19 guidelines) |
| *Specific guidelines for certain populations* | High risk groups: Guidelines mentioned cognitive impairments and psychotic disorders as potential populations with a higher risk when using telehealth.  1) Some guidelines suggested it may be preferable to exclude these from e-mental health interventions, but one set of guidelines noted that there is no concrete evidence indicating which populations may benefit most or may be harmed by psychological therapy delivered via videoconferencing.  2) No recommendations were made regarding adaption for these groups.  Young people 1) Six guidelines discussed appropriateness of e-mental health services for young people. 42% (8/19 guidelines) highlighted importance of explicitly checking age in young people and their consent as they can appear highly adult due to high computer literacy.  2) All guidelines noted that the requirement for parental consent should remain for e-intervention.  3) Guidelines did not discuss tailored strategies for young people. |
